# MOLAR: MRI-based Opportunistic Localization and Recognition of teeth

**DOI:** 10.64898/2026.09.18.26363455

**Authors:** Roger D. Newman-Norlund, Raghav Pallapothu, Santosh Kudaravalli, Jeet Sridhar, Pranesh Rajesh Kannan, Sriya Pallapothu

## Abstract

Tooth loss is associated with cognitive decline, yet dental status is rarely available in large neuroimaging cohorts, although routine T1-weighted (T1w) brain MRI captures the dentition. We present MOLAR (MRI-based Opportunistic Localization And Recognition of teeth), an open pipeline that isolates the dental region (the “bite box”) from T1w brain MRI and counts the teeth, without additional scanning or commercial software. Three raters annotated the usable OASIS-3 bite boxes (1,261 rated by all three; 63,829 point markers; count intraclass correlation coefficient, ICC(2,1) = 0.909). We reformulate counting from point labels as instance segmentation: each tooth becomes a compact capsule whose in-plane footprint adapts to voxel adjacency, so a standard three-dimensional nnU-Net counts teeth as connected components. On held-out scans the model detected teeth more consistently than raters agreed with one another (F1 0.873 against 0.787) and, after leave-one-fold-out threshold calibration, counted with mean absolute error 1.96 teeth, recovering 86% of the gap between an uninformed floor (3.57) and the inter-rater ceiling (1.70). A second network replaces registration by predicting the dental region, changing counts by 1.06 teeth, less than two raters disagree (1.58), whereas classical registration failed on 18% of scans. In an independent cohort (Aging Brain Cohort; n = 233; different scanner and raters), automated counts reproduced the manual association with the Montreal Cognitive Assessment (MoCA; Pearson r = +0.372 against +0.414, both p < 0.001), and after adjustment for age, sex and race the association was equivalent across counting methods. The software will be released open source.

**Graphical Abstract:** 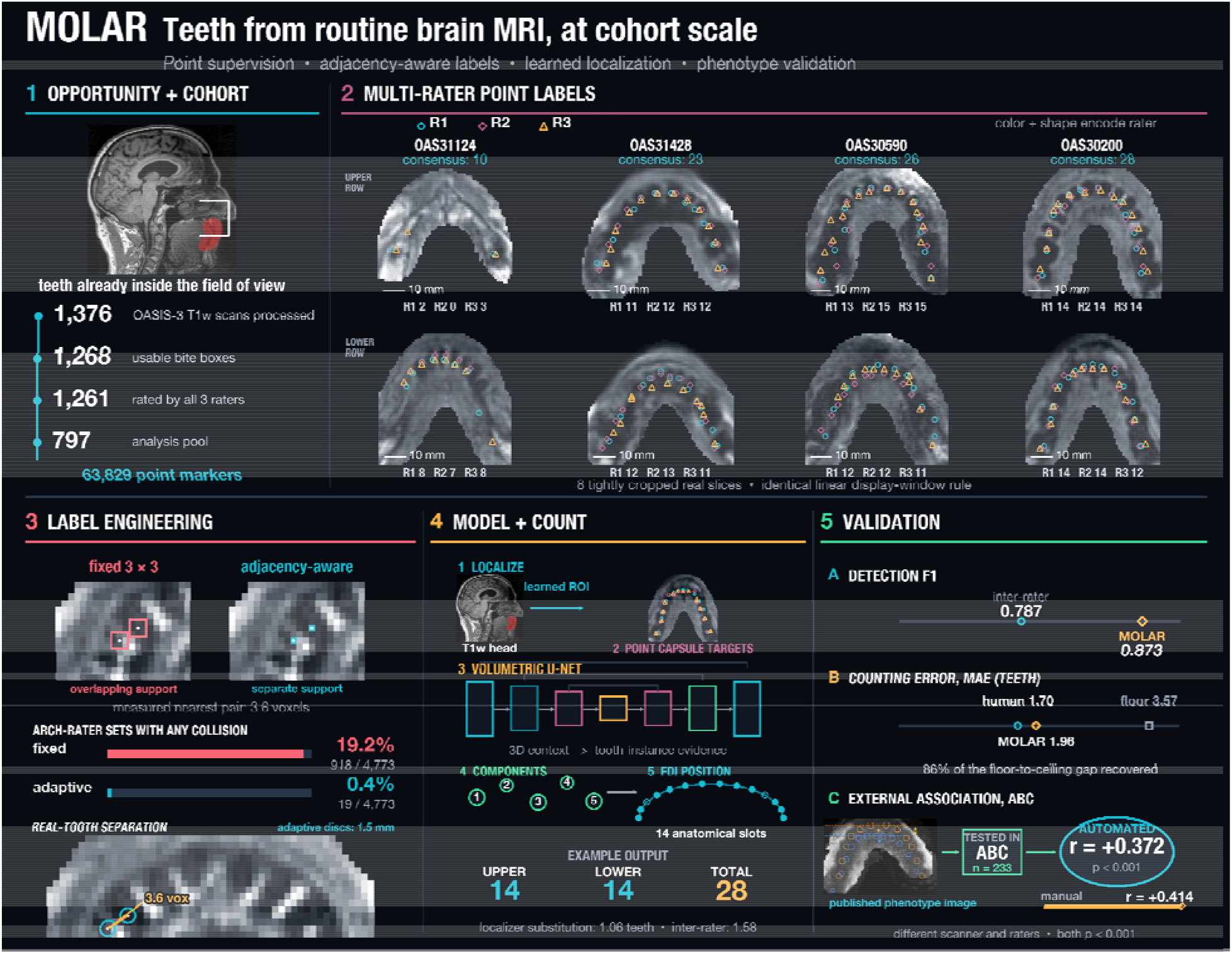

## 1. Introduction

Structural T1-weighted MRI is among the most widely shared data types in biomedical science, with open repositories distributing tens of thousands of whole-head acquisitions (LaMontagne et al., 2019; Jack et al., 2008; Van Essen et al., 2013; Sudlow et al., 2015; Bookheimer et al. 2019). Oral health has emerged as a consistent correlate of cognitive status and brain aging: tooth loss, periodontal disease and impaired masticatory function have each been associated with poorer cognition, accelerated decline and elevated dementia risk (Qi et al. 2021; Fang et al. 2018; Cerutti-Kopplin et al. 2016; Dintica et al. 2018), through mechanisms spanning neuroinflammation, oral microbial translocation, cerebrovascular injury and loss of masticatory sensory input (Weijenberg et al., 2011; Dominy et al., 2019; Kamer et al., 2015; Merchant et al., 2023; Beydoun et al. 2020). Because brain-optimized T1 acquisitions routinely extend inferiorly through the maxilla and mandible, the dentition is visible in a large fraction of archived scans, but no means of reading that information currently exists. Establishing dental status has required either a separate examination, which these studies were not designed to collect and cannot obtain retrospectively, or manual inspection of each volume by a trained rater, which is not standardized across observers and scales linearly with cohort size.

Tooth loss is irreversible, so the count accumulates a lifetime of oral disease rather than reflecting current activity. Thus, among measures of oral health, the number of remaining teeth is the most stable and most comparable across studies. It is the measure most consistently reported in epidemiological work relating oral status to cognition, requires no clinical judgment of disease activity, and is interpretable across health systems and decades (Thomson and Barak 2021; Qi et al. 2021); it also has functional meaning: teeth chew only in opposing upper and lower pairs, so the number remaining sets an upper bound on how many occluding pairs a person has, and thereby on chewing capacity and diet (Weijenberg et al., 2011). In our own work, manually counted MRI-derived tooth count correlated with Montreal Cognitive Assessment (Nasreddine et al. 2005) performance in the University of South Carolina Aging Brain Cohort after adjustment for age, sex and race, and improved identification of participants meeting criteria for clinically meaningful impairment (Newman-Norlund et al. 2024), establishing the association between tooth count and cognition that the present study uses as its external benchmark.

Large repositories are mined intensively, and almost exclusively, for brain-derived phenotypes: cortical thickness, subcortical volumes, white-matter hyperintensity load and brain-age estimates (Fischl 2012; Alfaro-Almagro et al. 2018; Cole and Franke, 2017; Bashyam et al. 2020; Wardlaw et al. 2013). Mature, freely distributed tools exist for each, and their availability is what made those measures standard (Smith 2002; Jenkinson et al. 2012; Billot et al. 2023; Henschel et al. 2020). No equivalent exists for the dentition. Automated dental analysis is well developed but operates on imaging designed for the purpose, where teeth are high-contrast and acquisition is standardized (Tuzoff et al. 2019; Cui et al. 2022; Schwendicke et al. 2019; Lee et al. 2018). Brain-optimized T1 MRI is a different regime: enamel and dentine are signal-void rather than hyperintense, contrast against bone and soft tissue is low, head position and field of view vary widely, and the dentition is frequently truncated. In this study we address that gap with three aims (**Figure 1**): (1) to localize the dental region automatically in whole-head T1w images, since neuroimaging templates are brain-centered and do not extend to the dentition; (2) to count teeth with a network trained only from point markers, since raters can mark tooth positions at scale but cannot trace boundaries at scale, and a count is recoverable from a segmentation only if adjacent teeth remain separate objects; and (3) to test whether the automated count reproduces, in an independent cohort, the association with cognition previously established from manual counts, since a measure can agree closely with manual counts on average while losing the between-subject variance that carries the association.

**Figure 1:**
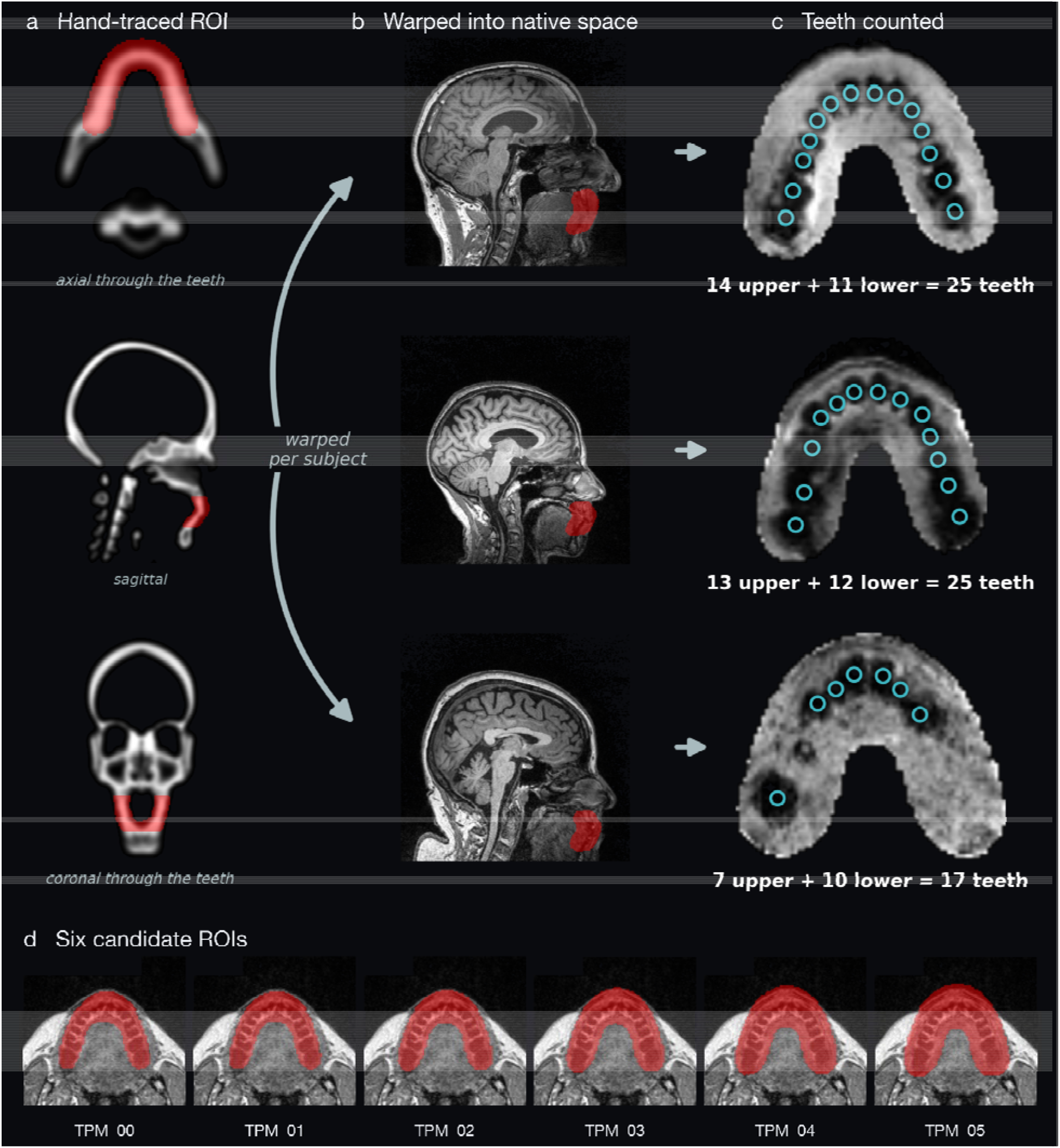
Bite-box extraction from whole-head T1w MRI. The only manual step is the one-time tracing in **(A)**; every subsequent step, including **(B)** and **(C)**, is automatic. **(A)** The anchor region of interest, a single expert tracing of all teeth drawn once on the bone compartment of an extended tissue-probability-map template in MNI space (Fonov et al., 2011), shown axial through the teeth, sagittal and coronal. **(B)** The tracing warped into three participants’ own native space on the whole-head T1w image; because the region is warped per subject, no two extracted boxes contain the same anatomical volume. **(C)** The bite box each warp yields, cut along the tooth row, with every tooth detected by the tooth model ringed and the upper and lower counts printed. Teeth are the dark structures: enamel and dentine are effectively MR-signal-void. **(D)** The six candidate regions of interest. TPM_00 is the tracing itself and TPM_01 to TPM_05 are progressive dilations that absorb registration error; all six are warped into native space and the smallest candidate passing quality control is selected per participant.

The methodological contributions are fourfold. (i) We cast counting from point annotations as instance segmentation, labeling each tooth as an anisotropic capsule whose in-plane footprint is set by voxel adjacency so that neighboring teeth remain separate connected components, which lets an unmodified nnU-Net count. (ii) We fuse markers from three raters by anchoring on the rater whose count matches the consensus, a rule that cannot chain across neighboring teeth, and show that three-rater supervision outperforms any single rater. (iii) We show that registration contributes only a binary region-of-interest mask to the pipeline and replace it with a learned localizer, evaluated by substitution into the full pipeline, with a paired usability classifier that allows the pipeline to decline scans. (iv) We validate the automated count as a phenotype in an independent cohort by testing whether it reproduces a published manual association with cognition, rather than by agreement alone.

## 2. Methods

### 2.1. Cohorts and bite-box generation

Development used 1,376 T1w scans from OASIS-3, one per participant (age 69.9 ± 9.7 years, 56% female), T1w being the most common structural acquisition and the contrast on which the segmentation priors are defined (LaMontagne et al. 2019; Marcus et al. 2007). External validation used 248 T1w scans from the University of South Carolina Aging Brain Cohort (ABC), a different study on a different scanner, counted by a different pair of raters using a per-quadrant protocol, with no overlap with the training data (Newman-Norlund et al. 2024). OASIS-3 participants gave informed consent under protocols approved by the Washington University Institutional Review Board; ABC participants gave written informed consent under a protocol approved by the University of South Carolina Institutional Review Board.

MOLAR isolates a bite box, the cropped sub-volume containing the dentition, from a whole-head T1w image. The method is anchored on a single expert-drawn region of interest (ROI) tracing all teeth, delineated once on the bone compartment of an extended tissue-probability-map template in MNI space (Fonov et al. 2011); five progressively dilated copies tolerate registration error, giving six candidate ROIs (**Figure 1d**). The tracing was drawn once, on the template, before the data were processed; it is never redrawn, and no human input is required when MOLAR is applied to a new scan. Each image is segmented with SPM12 priors (Ashburner and Friston 2005), the template bone compartment is normalized to the subject’s own, the transform is applied to the candidate ROIs, and the image is resliced to each native ROI grid and cropped to the bounding box of non-zero voxels (**Supplementary Methods S1**). Every bite box is scored on in-ROI volume, in-ROI signal fraction, contact with the field-of-view boundary and the number of large low-signal clusters, and assigned ok, review or fail with a reason.

### 2.2. Ground-truth tooth labeling

Three raters independently annotated every quality-control-passing bite box using a purpose-built browser tool built on the NiiVue WebGL NIfTI viewer (Hanayik et al. 2026; **Figure 2**, interface in **Figure S1**). Each rater assigned a usability rating (clear, partial or not usable) and placed one marker on each visible tooth, separately for the upper and lower arches, blinded to the other raters and to the automated heuristic. Reliability was quantified for total, upper and lower counts (ICC(2,1), mean absolute difference) and for usability (Cohen’s κ); because tooth visualization on brain-optimized T1w MRI is intrinsically variable, this agreement defines the human ceiling. Ground truth is derived by majority rather than by adjudication: the consensus count is the median of the three raters, and a tooth enters the reference set when at least two raters marked it within 3 mm in plane. The model was developed on the scans that all three raters rated clear or partial, that no rater flagged for metal artifact or a faulty bite box, and whose consensus count was nonzero (n = 797 of 1,261).

**Figure 2:**
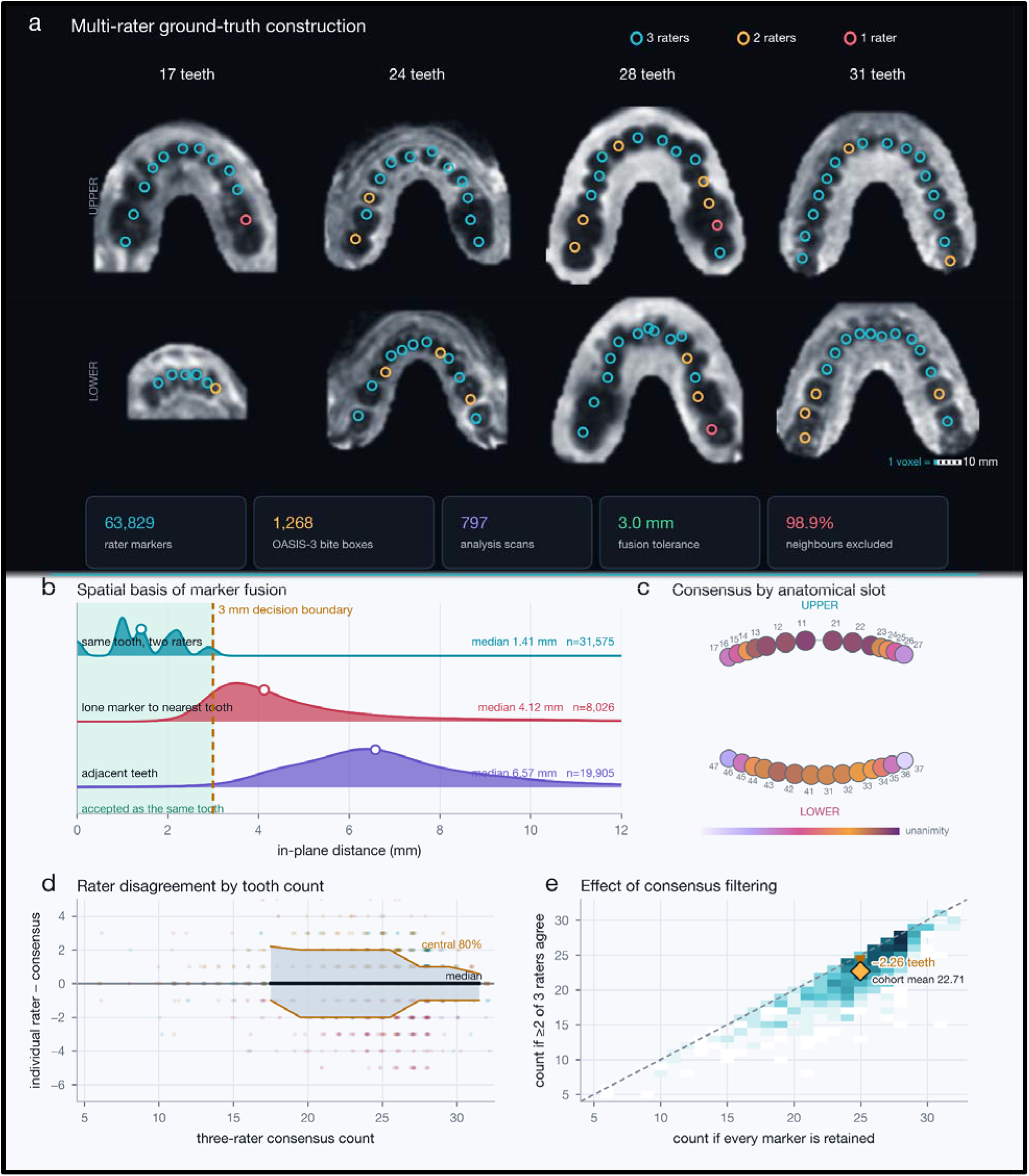
Ground truth annotation and the 3 mm matching rule. **(A)** Bite boxes from four OASIS-3 participants spanning the range of retained dentition, from 17 to 31 teeth by three-rater consensus; upper arch above, lower arch below, showing in radiological convention (the participant’s right on the image left). Each panel is a single axial slice and not a projection, because each rater annotated a given jaw on essentially one slice and different raters chose different slices. Marker coordinates are the annotators’ own and are never altered; only the slice on which they are displayed is chosen, from the two slices either side of the annotation slice, so that the rings sit deepest in the tooth void. One ring is drawn per tooth the fusion rule retains, at the anchoring rater’s marker position, colored by how many of the three raters independently marked it. **(B)** The 3 mm matching tolerance in context: the joint distribution of the offset between two raters marking the same tooth, and the cumulative distribution of that offset, of the distance from a lone marker to the nearest corroborated tooth, and of the distance to the nearest other tooth in the arch. Every rater offset falls inside 3 mm and 98.9% of neighboring teeth lie beyond it. **(C)** Slot occupancy and three-rater unanimity by FDI tooth position, pooled across the cohort rather than shown for one participant: disc area gives the share of participants in whom that tooth position is filled, and disc color is a continuous scale giving, among those participants, the share of teeth at that position marked by all three raters. **(D)** Deviation of an individual rater’s count from the three-rater consensus, with the 10th and 90th centiles traced. **(E)** Tooth count under the two label definitions, majority against every marker, whose cohort means are 22.71 and 24.98 teeth. Panels (B) to (E) are computed on the 797-scan analysis pool.

### 2.3. Counting as instance segmentation

The annotations are one point per tooth, so the supervision is spatial rather than scalar. Regressing a single count would discard 63,829 point labels and return a number that cannot be audited against the image, so we formulated the task as instance detection and made the count a property of the detected set. Unsupervised alternatives fail: teeth are reliably dark (**Figure S2**), but darkness is not discriminative, 98-99% of sub-threshold voxels within a bite box forming one connected component spanning the teeth, oral cavity and airway (**Supplementary Methods S2).**

Semantic segmentation networks assign classes, not instances. Labeling all teeth as one class makes neighbors merge into an uncountable blob; labeling 32 FDI classes requires per-tooth identity that no rater provided. We resolve this geometrically: each tooth is labeled as a small capsule centered on its consensus marker, a 1.5 mm in-plane disc extended ±3 mm in z, class 1 upper and class 2 lower, so that tooth count is the number of connected components per class. Of the 20,917 fused markers, 19,905 fall in arches with two or more teeth and enter the spacing analysis. Measured arch geometry licenses the reformulation, since across 19,905 teeth the median in-plane nearest-neighbor spacing is 6.57 mm (1st percentile 3.00 mm), so capsules remain separate (**Figure S3**). The capsule is anisotropic by design, teeth being columns in z, so extending in z raises the foreground fraction about ninefold without changing the in-plane collision rate counting depends on.

Component separation is governed by voxel adjacency rather than Euclidean distance: under 6-connectivity two full 3 × 3 discs remain distinct only at a Chebyshev separation of 4, not 3. Each tooth’s in-plane footprint is therefore set adaptively, the full disc where the nearest neighbor is at least four voxels away and a single-voxel column otherwise (**Figure 3**), reducing the share of arch-by-rater label sets containing any merged pair from 19.2% (918 of 4,773) to 0.4% (19 of 4,773). Because raters mark the same tooth at slightly different positions and on their own chosen slice, markers are fused before labeling by anchoring on the rater whose count equals the consensus and refining each position from the other raters’ nearest markers within 3 mm, a rule that cannot chain across neighbors as single-linkage clustering does (**Supplementary Methods S3).**

**Figure 3:**
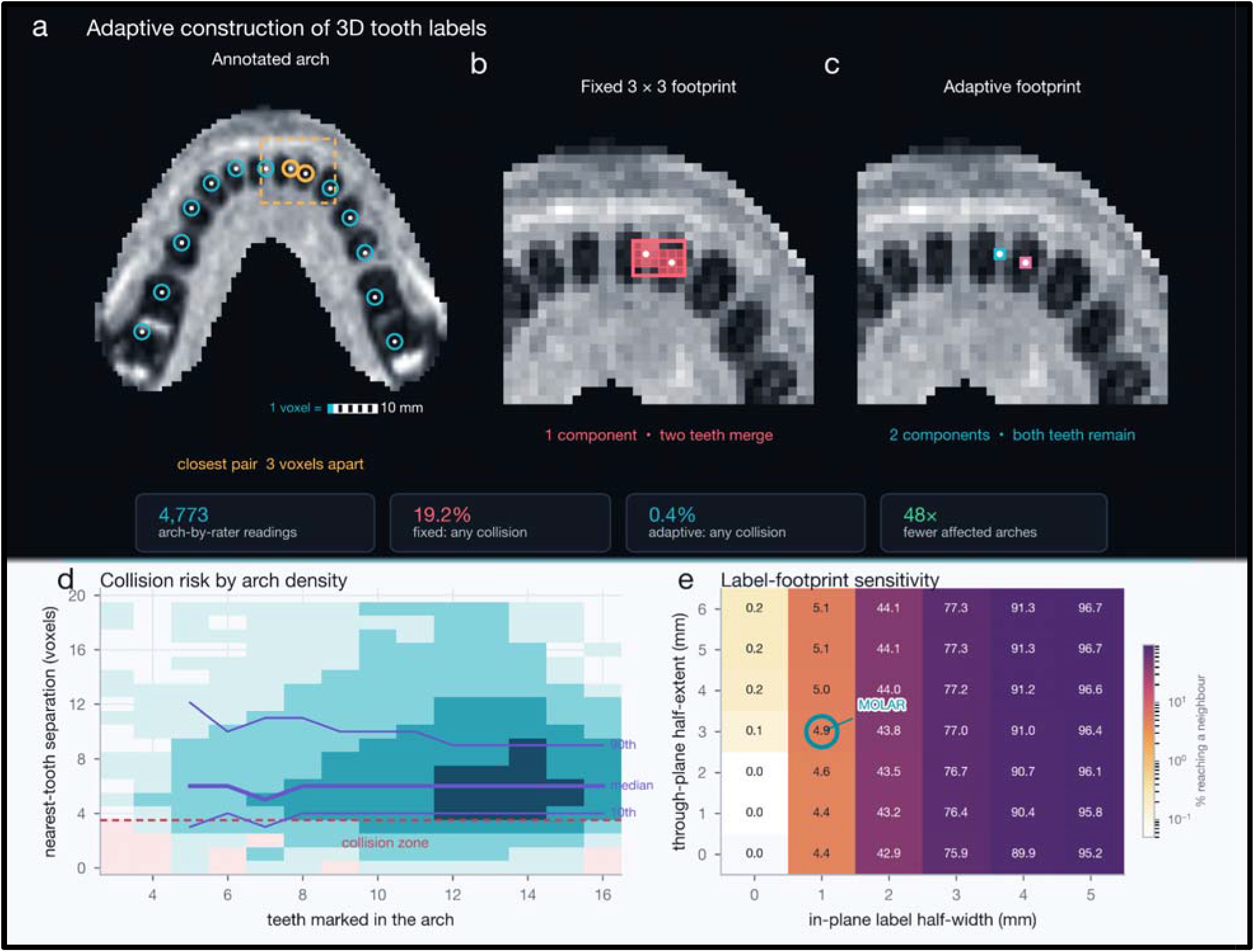
The adaptive capsule footprint keeps neighboring teeth separate. **(A)** One rated lower arch, minimum-intensity projection through the marker slab, showing all of that rater’s markers and the capsules built from them: a 1.5 mm in-plane disc extended ±3 mm through the plane. Amber marks the closest pair of teeth in this arch and the dashed box is the field enlarged in **(B)** and **(C). (B, C)** That pair at voxel resolution, both panels on one intensity window taken from the whole bite box so that the two rules can be compared directly. Under the fixed rule **(B)** every tooth receives the same 3 × 3 disc; at the Chebyshev separation of 3 voxels seen in this pair the two discs become face-adjacent, 6-connected component labeling returns one component, and two teeth are counted as one. Under the adaptive rule **(C)** a tooth whose nearest neighbor is closer than 4 voxels is stamped as a single-voxel column instead, and component labeling returns two components. **(D)** Merge risk does not follow tooth count: 63,829 rated teeth in 4,773 arch-by-rater sets, binned by the number of teeth the rater placed in that arch against Chebyshev separation to the nearest tooth, with the 10th, 50th and 90th centiles traced. **(E)** Sensitivity of label merging to the capsule’s in-plane and through-plane half-extents: the proportion of labels that reach a neighbor, with the selected MOLAR footprint highlighted. Across the 4,773 arch-by-rater label sets, the adaptive footprint reduces the share containing any merge from 19.2% (918) to 0.4% (19) and keeps 99.7% of teeth separate.

### 2.4. Model and derived tooth identity

We use a three-dimensional U-Net (Ronneberger et al., 2015; Milletari et al., 2016) configured automatically by nnU-Net (Isensee et al., 2021), chosen after considering the alternatives in **Table S1**; self-configuration removes choices that would otherwise be tuned on a small sample. We compare the default encoder against the larger ResEnc-M preset and against a two-dimensional configuration, under five-fold cross-validation stratified by count and field strength with one scan per participant. Raters did not label tooth identity, so FDI codes are derived rather than supervised: ordering detections by left-right coordinate recovers the anatomical sequence, and observed teeth are assigned to canonical slots by a monotone alignment permitting empty slots, so a partial dentition is numbered with gaps rather than renumbered consecutively (**Figure 4; Supplementary Methods S4).**

**Figure 4:**
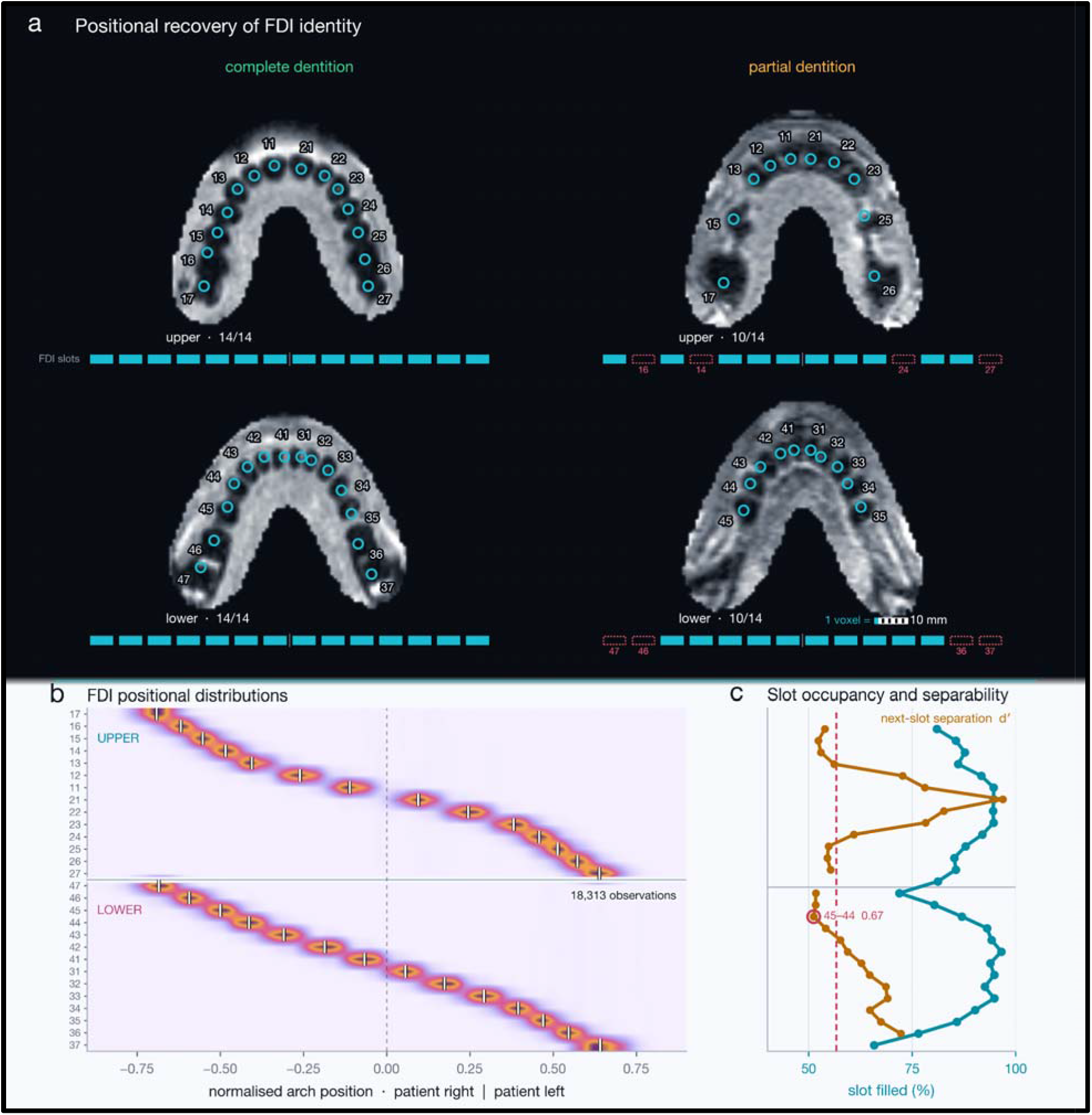
Tooth identity assigned from arch position without supervision. FDI codes are derived rather than supervised: detections are ordered along the arch and assigned to canonical slots by a monotone dynamic-programming alignment that permits empty slots, so a partial dentition is numbered with explicit gaps rather than renumbered consecutively. **(A)** Complete (left) and partial (right) upper and lower arches on real bite boxes, with the raters’ own marker coordinates and the derived FDI codes; the rail beneath each arch shows its 14 canonical slots, filled where a tooth is assigned and outlined in coral where the slot is empty. The partial case is missing 16, 14, 24 and 27 above and 47, 46, 36 and 37 below. No rater ever labeled a tooth number: ordering markers along the arch and aligning them to the canonical slots, allowing gaps, recovers the codes and names which teeth are absent. **(B)** Density of normalized arch position for each of the 28 FDI slots across 18,313 placed teeth, with slot medians as white ticks; the canonical slot positions were estimated from the 852 complete upper and 637 complete lower arch ratings. **(C)** Slot occupancy (share of arches in which the slot is filled) and next-slot separability d′ by FDI position; the highlighted pair (45 to 44, d′ = 0.67) is the most overlapping adjacent pair. Laterality is taken from the image affine and requires independent confirmation before FDI codes are released for use.

### 2.5. Learned localization of the dental region

The reference ROIs are produced by registration, the one part of the pipeline requiring commercial-adjacent tooling, which raises a methodological question: is registration doing work a network could do directly? Reimplementing the post-normalization stages in Python and supplying them the reference ROI reproduced the reference bite box bit-for-bit on every case tested, establishing that registration’s entire contribution to inference is the production of a single binary ROI mask. We therefore trained a localizer, a three-dimensional U-Net under the identical self-configuration procedure, taking the whole head at 2 mm and predicting the dilation-05 ROI directly (n = 1,268 after excluding QC failures as label noise). Because a localizer can misplace the region or place a correct region on the wrong grid, we evaluate by substitution into the pipeline while holding the tooth model fixed (**Table 1**). Accuracy is additionally reported as centroid displacement rather than as Dice, since an overlap coefficient computed against a dilated template tracing would reward agreement with an arbitrary edge.

A tool intended to run unsupervised must be able to decline. The three-rater majority verdict on whether the dentition was countable becomes a binary label, and a compact three-dimensional convolutional network (roughly 2 × 10□ parameters) is trained on the bite box alone to predict it under the same cross-validation. The binary target is used rather than the three-level grade, agreement on which is only moderate (κ 0.55-0.63). The classifier receives only the image, the pipeline’s own quality-control metrics being withheld as the baseline it must beat. Counts are scored by mean absolute error (MAE), bias and exact, within-1 and within-2 agreement; detections by in-plane precision, recall and F1 at 3 mm and 4 mm tolerance, scored identically to rater-versus-rater agreement so the two are directly comparable. Dice is deliberately not the primary metric (Maier-Hein et al. 2024): the targets are approximately 61-voxel capsules, so a one-voxel center offset collapses Dice while leaving the count correct.

### 2.6. External validation and statistical analysis

The primary external-validation outcome was the Montreal Cognitive Assessment (MoCA; Nasreddine et al., 2005), the outcome of the published manual analysis in this cohort (Newman-Norlund et al., 2024). The secondary outcome, also examined in that analysis, was the brain-age gap (BAG), defined as MRI-predicted brain age minus chronological age, so that a positive BAG indicates a brain that appears older than expected for the participant’s age (Cole and Franke, 2017). BAG was computed with two independent estimators: brainageR (Cole, 2019), the estimator used in the published manual analysis (n = 231), and DeepBrainNet (Bashyam et al., 2020), a deep network trained on 14,468 individuals, available for a subset (n = 141) and included to test whether any association depends on the choice of estimator. Associations between tooth count and each outcome are reported as Pearson correlations and as partial correlations from a linear model containing age, sex and race, the covariate set of the published manual analysis. The derived occlusal measures were exploratory and are Holm-corrected for the number of measures tested. Tests are two-sided with α = 0.05.

## 3. Results

### 3.1. Cohort processing

All 1,376 scans processed in 8.4 h with two hard errors; automated quality control classified 1,070 (77.8%) as ok, 198 (14.4%) as review and 106 (7.7%) as fail, so 1,268 scans (92.2%) yielded a usable bite box, the dominant exclusions being field-of-view truncation and low in-ROI signal (**Figure S4**). Every stage downstream of registration reproduced the reference output bit-for-bit, so differences between localization arms are attributable to the region of interest, not numerical drift.

### 3.2. Inter-rater agreement and tooth-model performance

Of the 1,268 usable bite boxes, 1,261 were annotated by every rater, yielding 63,829 tooth markers. Count agreement was high (Shrout and Fleiss 1979; Koo and Li 2016): ICC(2,1) = 0.909 total, 0.904 lower, 0.860 upper. Categorical judgments were less consistent, usability agreeing exactly across all three raters in 55.4% of cases (κ 0.55-0.63) (Landis and Koch 1977), though collapsed to the binary distinction that gates analysis, agreement was strong (κ 0.76-0.90). Restricted to the analysis pool (n = 797), rater-versus-rater count MAE was 1.36-1.91 teeth (mean 1.70) with in-plane detection F1 of 0.787 at 3 mm and 0.872 at 4 mm. These values, not the wider agreement over all 1,261 scans (MAE 2.0-2.8, inflated by unusable and edentulous cases the model never sees), are the ceiling against which the model is judged. Three raters also outperform any one rater (MAE 1.702 from a single rater against 1.513 from the mean of two), and the raters differ systematically (biases +0.53, +0.34 and −0.88 teeth), so a model trained on one rater inherits that rater’s offset. A constant predictor achieved MAE 3.57 on the analysis pool, and a naive dark-blob detector far worse (MAE 18.71).

Across every configuration the model localized individual teeth more consistently than the raters agreed with one another (**Figure 5**), reaching in-plane detection F1 of 0.873 at 3 mm against a rater ceiling of 0.787, and 0.892 against 0.872 at 4 mm. Detection and counting therefore dissociate: finding the teeth is the easier half, and the residual error lies in deciding how many objects the detected evidence represents. The default three-dimensional configuration outperformed both the larger ResEnc-M preset and a two-dimensional configuration (MAE 2.33, 2.46 and 4.41; **Table S2**), confirming that three-dimensional context is needed for a structure elongated in z. Training on the three-rater consensus also beat training on any single rater, scored against every rater in turn so that no arm is graded by the rater that trained it (**Table S3).**

**Figure 5:**
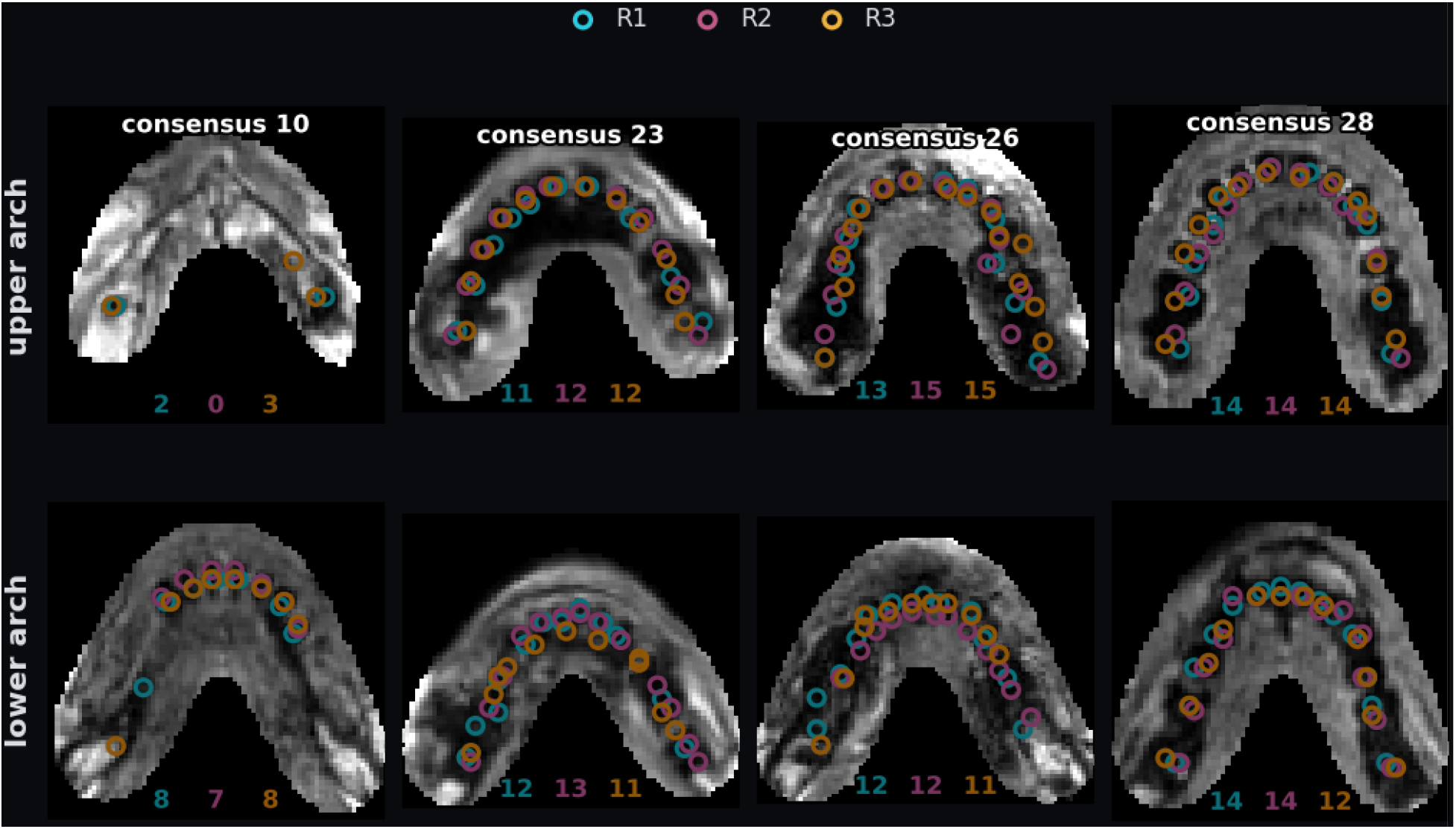
Inter-rater variability in tooth marking. Four OASIS-3 participants spanning the range of retained dentition, upper arch above and lower arch below, with each rater’s markers drawn in that rater’s own color on the slice that rater used. The number above each column is the participant’s whole-mouth three-rater consensus count and the three numbers below each arch are the individual rater counts for that arch. Agreement is close where the dentition is intact and degrades where it is not: in the leftmost participant the upper arch draws counts of 2, 0 and 3 against a whole-mouth consensus of 10, while in the rightmost every rater returns 14 above. This is the disagreement every model number in the paper is judged against, and it is why a tooth enters the reference set only when at least two raters mark it within 3 mm in plane. Teeth are the dark voids, enamel giving no MR signal.

Calibration, not architecture or label source, is the dominant error source. At the default decision rule the model undercounted the target it was trained on by 1.59 teeth, a recall deficit produced by training with about 0.7% foreground, the network under-segmenting to protect precision (0.916 against recall 0.826); it is not caused by adjacent detections merging, since predicted components were 0.77 times the size of label components and eroding them made counting worse. Lowering the tooth-probability threshold removes most of the bias. The threshold is selected leave-one-fold-out, so it is never tuned on the data it is scored on, and the same value (0.02) won on all five folds, giving MAE 1.96 teeth (SD 0.10) with a residual bias of −0.37, against arg-max values of 2.39 and −1.48. Against an inter-rater ceiling of 1.70 and an uninformed floor of 3.57, the calibrated model recovers 86% of that distance, and training for 1,000 epochs rather than 250 changed nothing, locating the remaining limit in the labels and the modality rather than in optimization. Bias must be assessed against the target a model was actually trained on, since scoring an all-marker model against the majority target produces a spurious appearance of robustness from two canceling offsets (**Supplementary Results S1).**

### 3.3. Contribution of registration and performance of the learned localizer

With the reference ROI supplied, the pure-Python implementation of every post-normalization stage reproduced its bite box bit-for-bit, and the output grid proved not to be a template-space reorientation but the subject’s own voxel lattice, x-flipped and resampled to 1 mm, differing only by a sub-voxel phase offset carrying no anatomical information (**Supplementary Results S3**). The question therefore reduces to whether the ROI mask can be predicted, and it largely can. Over all 254 held-out scans, median bite-box centroid displacement was 1.37 mm (66.5% within 2 mm) but 11.8% exceeded 10 mm (**Figure S5**). That aggregate pools scans no one would analyze: stratified by whether a scan entered the analysis pool, the failure rate is 1.3% inside the pool (median error 1.16 mm) and 29.5% outside it. The failing scans are field-of-view truncated (44% edge-clipped against 1.8%) with reduced in-ROI signal, which is why reviewers excluded them: the dentition is not fully present in the image.

Failure rate tracks input field-of-view shape (**Figure S5**), but shape marks truncated scans rather than causing failure, since rebuilding the training set in a fixed 128³ 2 mm cube left it unchanged. The controllable driver is acquisition obliquity, the scans the localizer mislocates being markedly more oblique than those it gets right (mean 15.2° against 9.7°; **Supplementary Results S2**), so rigid alignment to a common frame before localization is the appropriate remedy and the one outstanding modeling item. Such failures can be flagged, though not from one prediction, since a wrong box has a normal ROI volume: combining the image-side quality control with disagreement between two independently trained localizers catches 73% of gross failures for a 9% false-alarm rate, cutting the residual failure rate among passing scans from 13.0% to 4.3% (**Supplementary Results S2**). The released tool therefore runs both localizers and reports their disagreement per scan.

Substituted into the full pipeline, learned localization cost approximately one tooth (**Table 2**, **Figure 6a–c, g**): replacing the registration grid alone changed the count by 0.43 teeth and predicting the ROI added a further 0.95, for a total of 1.06 teeth with a bias of −0.07. Two raters disagree by 1.58 teeth on the same scans, so the substitution perturbs the count less than raters disagree and does not shift the cohort mean, the property that matters downstream. Classical registration is not a viable substitute: a SimpleITK implementation warping the template directly to the subject with Mattes mutual information (Lowekamp et al. 2013; Jenkinson et al. 2002; Avants et al. 2011) produced a grossly wrong box on 29 of 158 scans (18%), against 1.3% for the localizer.

**Figure 6:**
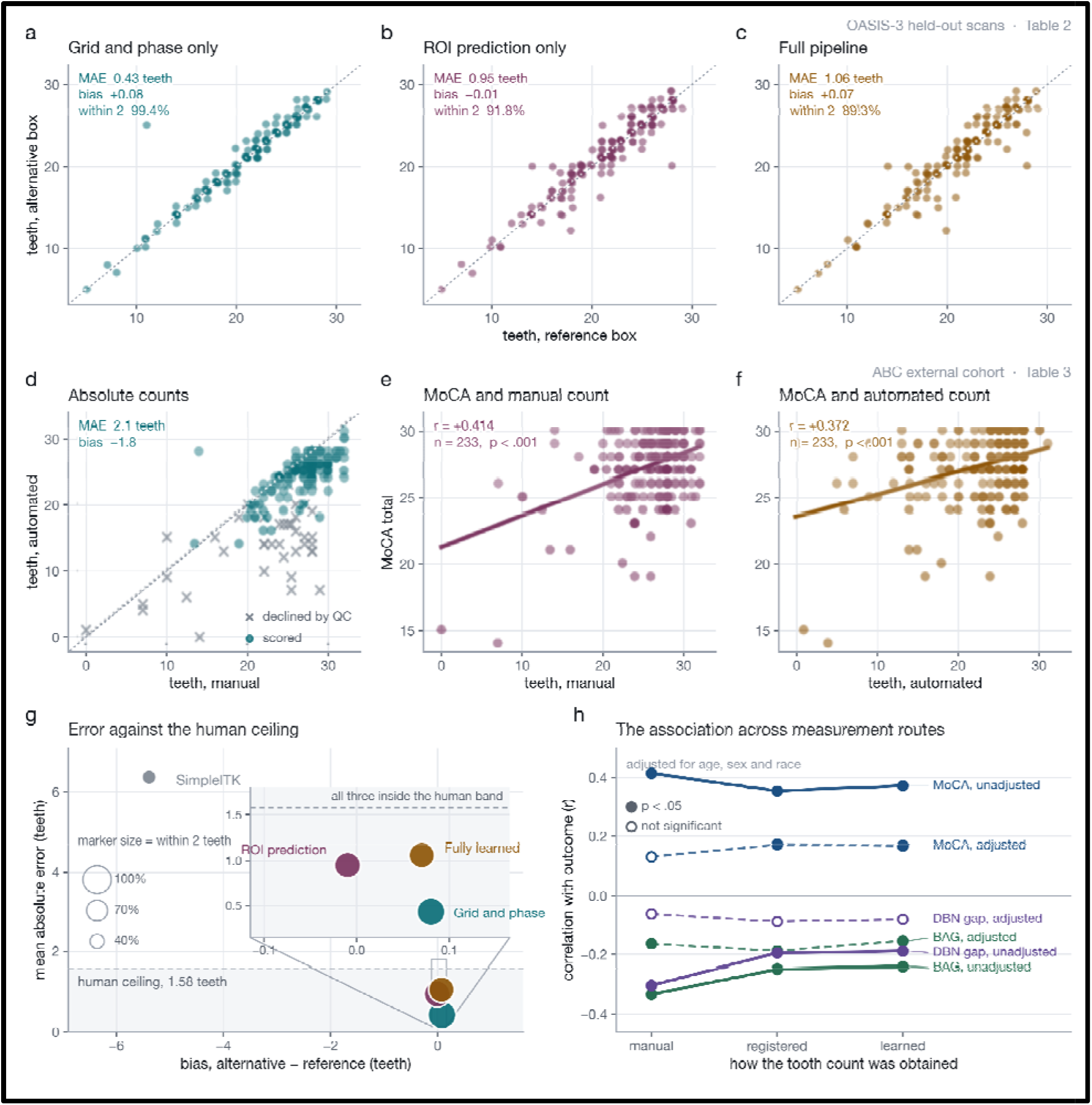
Localization costs and whether automated count carries the association. **(A-C)** Tooth counts from bite boxes built three ways on the OASIS-3 held-out scans, with the tooth model held fixed so that every difference is caused by box construction alone (**Table 2**). **(A)** The reference region on a canonical grid, isolating the effect of abandoning the registration grid’s arbitrary sub-voxel phase offset. **(B)** A predicted region on the same grid, isolating localizer error. **(C)** The fully learned pipeline. The dotted line is identity. **(D-F)** The same pipeline on the ABC cohort (**Table 3**). **(D)** Automated against manual counts, with scans the pipeline declines to score shown as crosses; every catastrophic disagreement is a declined scan, the cases with 20 or more teeth counted manually and fewer than ten found automatically all falling below the usability gate. On the scans it does score the automated count sits 1.8 teeth below the manual count, an offset that follows from the conservative label definition. **(E, F)** The tooth-cognition association measured with manual and with fully automated counts in the same 233 ABC participants. **(G)** All four localization routes in the bias by mean-absolute-error plane, marker area giving the share of scans within two teeth of the reference. The shaded band is the 1.58-tooth disagreement between two human raters; the inset resolves the three learned arms, all of which fall inside it, while a classical SimpleITK registration does not. **(H)** The correlation with each outcome under the three measurement routes, unadjusted and adjusted for age, sex and race, filled markers marking p < 0.05. What survives adjustment is equivalent across routes, which is the property that matters for use as a phenotype.

**Figure 7:**
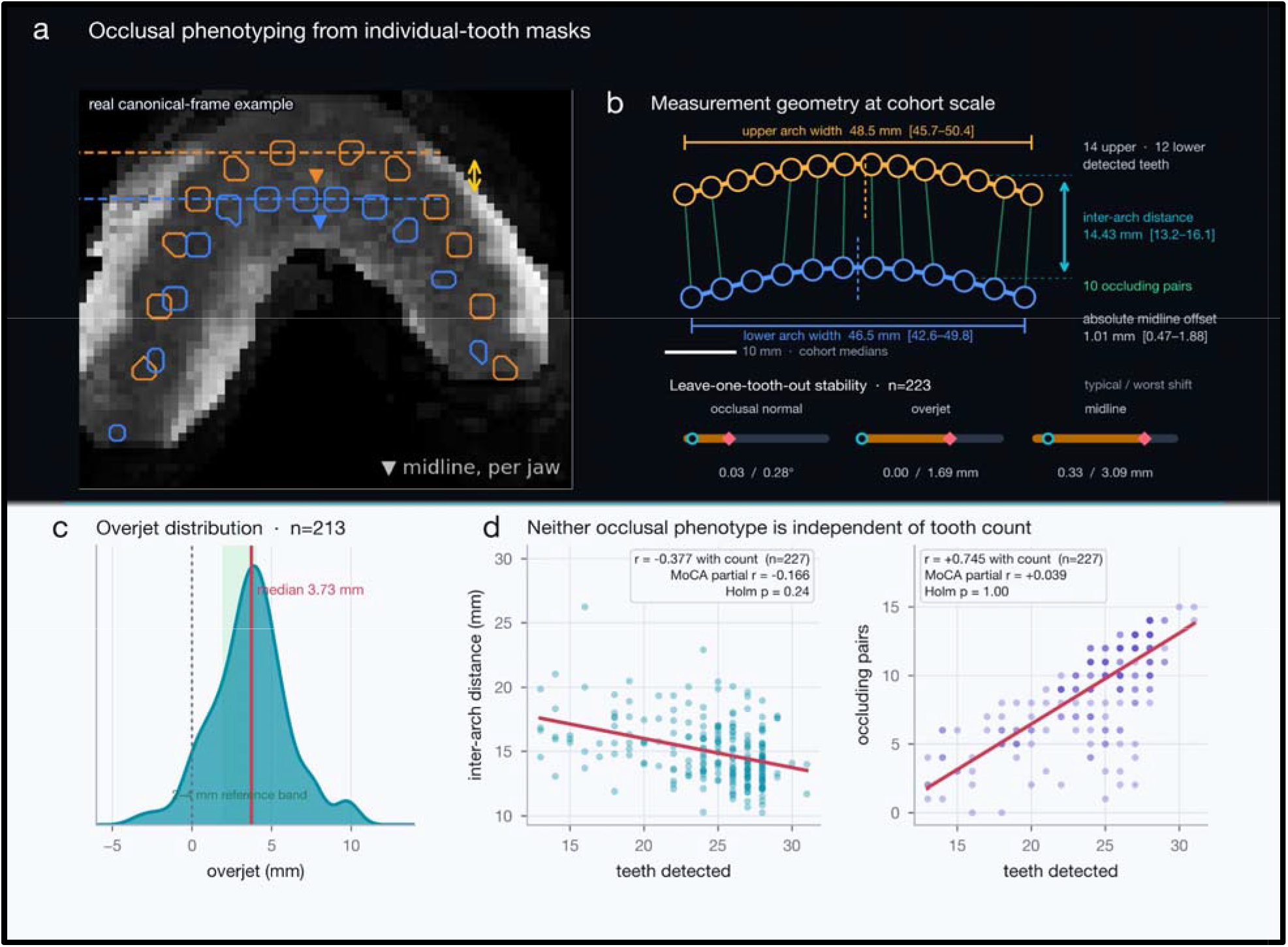
Occlusal phenotyping from the same bite boxes. **a,** Canonical-frame upper (amber) and lower (blue) tooth masks, each detected tooth outlined on a minimum-intensity projection viewed down the occlusal normal, so the whole arch appears in one image with teeth as dark voids. **b,** Measurement geometry drawn at cohort scale (n = 213 with a full measure set): arch width (upper 48.5 mm [45.7–50.4]; lower 46.5 mm [42.6–49.8]), centroid-to-centroid inter-arch distance (14.43 mm [13.2–16.1]), absolute jaw-specific midline offset (1.01 mm [0.47–1.88]), and the median dentition of 14 upper and 12 lower detected teeth forming 10 occluding pairs. Every dimension and count is a cohort median of a measured quantity, drawn to the millimeter scale shown; only the arch curvature is a drawing convention. The band beneath reports leave-one-tooth-out stability in 223 participants: typical and worst-case changes of 0.03°/0.28° for the occlusal normal, 0.00/1.69 mm for overjet and 0.33/3.09 mm for midline position. **c,** Overjet in the complete subset (n = 213; median 3.73 mm), with the range a clinical examination would call normal shaded. d, Both phenotypes against tooth count (n = 227): inter-arch distance falls as teeth are retained (r = −0.377) and occluding pairs rise with them (r = +0.745). Neither association with MoCA survives Holm correction (inter-arch r = −0.166, Holm p = 0.24; occluding pairs r = +0.039, Holm p = 1.00). These are exploratory derived phenotypes and have not been validated against a dental examination.

### 3.4. Automated usability prediction and external validation

The four quality-control metrics the pipeline already computes carry almost no information about whether a human would judge a scan countable (area under the curve 0.636), having been built to catch gross localization failures instead. A classifier reading the bite box itself answers that question: cross-validated on the 1,268 scans with a usability majority it reaches AUC 0.968 and agrees with the rater majority on 90% of scans in both directions, against 83.5% unanimity among the raters themselves. The pipeline therefore does not need a human to decide which scans it can handle, which is what allows it to be applied to a whole archive rather than a curated subset.

Bite-box extraction succeeded on essentially all 248 ABC scans (99.6% usable), against 86% failure on an earlier defaced release of the same scans (Bischoff-Grethe et al. 2007; Schwarz et al. 2019), confirming that face-preserving reconstructions are a hard requirement for dental measurement. Excluding eight scans on which localization failed outright and six identifiers sharing three byte-identical source images with irreconcilable demographics (**Supplementary Results S5)** leaves 234 analyzed. Absolute counts are offset, expectedly, because the model predicts the conservative OASIS majority label (training mean 22.8 teeth) whereas the ABC raters’ convention gives a mean of 26.2 here. The automated mean of 23.2 is close to what the label definition predicts, so the bias (−2.91 teeth, MAE 3.16) reflects a difference in what is counted rather than a failure to count, narrowing to −1.85 teeth (MAE 2.09) on the scans the pipeline judges countable. The ABC raters also agreed with each other far more closely than the OASIS raters did (0.72 teeth against 1.58), so ABC is a stricter comparison than the training cohort.

Fitness for use as a phenotype depends on whether the automated measure reproduces the association obtained from the manual counts, since a constant offset leaves a correlation unchanged. The association is reproduced (**Table 3**, **Figure 6d–f, h**). Unadjusted, tooth count and MoCA correlated at Pearson r = +0.414 for manual and +0.372 for fully automated counts (n = 233, both p < 0.001). Against the model used in the published manual analysis, which adjusted for age, sex and race, the partial correlation was +0.134 for manual counts (p = 0.067), +0.174 for registration-derived automated counts (p = 0.017) and +0.170 for the fully learned pipeline (p = 0.020). The manual counts are those of the published analysis, not a recount; the estimate differs from the published one-tailed partial r(208) = 0.233 (Newman-Norlund et al., 2024) because this analysis uses a different subsample, the 192 participants with the full covariate set among the 233 scans remaining after localization failures and duplicate identifiers were excluded, and reports two-tailed tests. Adjustment reduces every estimate by half or more, but residual association is equivalent across the three measurement routes, and the automated measures reach conventional significance in this sample where the manual counts do not. For the secondary outcome, BAG (Section 2.6), the age-adjusted association was also equivalent across routes (partial r = −0.164 manual, −0.188 registration-derived and −0.154 learned; n = 191, all p < 0.05), so participants with more teeth had brains that appeared younger for their age regardless of how teeth were counted. With the DeepBrainNet estimator no route reached significance after adjustment (n = 104), which, given the smaller sample, we treat as inconclusive rather than as evidence against the association.

The usability gate is a reporting device rather than a cohort filter. To test whether scan quality explains the association, we repeated the analysis restricted to the scans the gate accepts, as a sensitivity analysis; usability was not entered as a covariate, because it is a consequence of the exposure (sparser dentitions produce less tooth-like bite boxes) rather than a confounder. Applied at its operating point, it retains 192 of 233 scans and removes the association in every arm, the manual counts included (adjusted r = +0.002 manual, +0.097 registration-derived, +0.079 learned; n = 159, the retained scans with the full covariate set), because unusability is not independent of what is being measured: declined scans average 22.3 teeth, against 27.0 for those retained (p = 6 × 10□□), so it discards the low end of the exposure and the resulting range restriction attenuates the correlation (**Supplementary Results S4**). The attenuation is equally severe for the manual counts, which identifies it as a property of the selection rather than of the automated measure, so the gate should flag images on which the tool is unreliable rather than select an analysis sample when tooth count is the exposure.

The analyses above used registration-derived bite boxes, which tests the tooth model externally but not the learned localizer. Repeated with every ABC bite box placed by the localizer, the pipeline produced a box for all 248 scans without failure and agreed with the ABC raters at least as closely as the registration pipeline (MAE 3.16 against 3.56), and the fully learned pipeline reproduced the tooth-cognition association at Pearson r = +0.372 (**p < 0.001**), so the choice of localization path does not determine whether the association is detected. Since the model segments teeth individually rather than regressing a number, the arrangement of the dentition can be measured at no extra cost: overjet, inter-arch distance, arch width and curvature, midline offset and the number of occluding pairs, read in a per-participant canonical dental frame. These measures are internally consistent and fall in the ranges a dental examination would give, but none has been compared against an examination of the same mouth, none is more predictive of cognition than the tooth count itself, and one candidate measure (overbite) is not recoverable from this modality at all. They are reported in full in the Supplementary material as exploratory outputs and form no part of the claims made for the method here.

## 4. Discussion

MOLAR makes the incidental dental content of brain MRI usable at scale, with no additional scanning, no commercial software and no manual annotation at inference time; given a T1 it returns a dental region, a per-tooth segmentation, a tooth count and quality flags. The measure is not a clinical tooth count and should not be reported as one: it is the number of teeth visible and corroborated on a brain-optimized image, and because the training label counts only teeth marked by at least two of three raters, cohort means sit several teeth below a clinical per-quadrant convention. That offset is stable, so it cancels in correlations in group comparisons, which is how the measure is intended to be used. The tooth loss, cognition, and brain health relationship is already established, including in this cohort. The contribution here is not the association but the replacement of manual counting with a measure applicable to cohorts of any size, and the validation is framed accordingly: the automated count reproduces the manual count-cognition association at p < 0.001 in an independent cohort with roughly 90% of the manual effect size. For the secondary brain-age outcome the age-adjusted automated estimate was 94% of the manual one, while the smaller DeepBrainNet subsample (n = 104) was underpowered for any route to reach significance. Automation creates a larger sample size, but not sensitivity per participant.

Three methodological points generalize beyond this pipeline. First, a legacy component may not be doing what its name implies: the normalization step here wrote onto the subject’s own voxel lattice rather than reorienting into template space, so an apparent dependence on a registration framework was in fact a dependence on a single binary mask, and auditing what it contributed is why a learned localizer could replace it at a cost of one tooth. Second, an intuitive substitute is not automatically a working one, classical registration failing on 18% of scans where the localizer failed on 1.3%. Third, aggregate failure rates can mislead: the localizer’s apparent 11.8% failure rate resolved into 1.3% on analyzable scans and 29.5% on scans raters had already rejected, and a compelling correlation between failure and acquisition geometry did not survive intervention.

Silent failure is the residual risk and is partially addressable, since flagging disagreement between two localizers with different input geometries, together with field-of-view and signal checks, catches 73% of gross failures at a 9% false-alarm rate. The remaining exposure is correlated blind spots between the two models, which is why the tool reports flags rather than silently discarding scans. Two alternative formulations were not evaluated, for reasons of scope rather than merit: purpose-built object detection (Baumgartner et al. 2021), and point supervision with a center-heatmap and offset field.

The principal limitation is that the measure is calibrated to a rater definition rather than to a dental examination. No participant in either cohort has a clinical dental record, so we can report how closely the automated count reproduces expert counting of the same images, the task it is designed to replace, but not its agreement with a mouth; anchoring against examination in a subsample remains the most valuable next step. Individual detections were scored against the raters’ markers rather than adjudicated one by one, so the reported precision is a lower bound, and counting remains distinguishable from human performance (MAE 1.96 against a 1.70 ceiling), although tooth localization already exceeds inter-rater consistency, which places the remaining gap in the count decision rather than in detection. Ground truth is tooth count, not per-tooth identity, so the FDI numbering derived from arch position is unvalidated, and defacing is disqualifying, since it removes the anatomy being measured. Finally, both cohorts are research cohorts acquired on a stable protocol, and that is the setting the method is suited to. A clinical archive, in which resolution, field of view, sequence and head placement all vary, is a different proposition: truncation removes teeth from the image rather than degrading them, so where it is not random with respect to the research question it becomes a bias rather than noise, and such an archive would require per-scan gating and probably retraining. Beyond examination anchoring, the natural extensions are richer dental phenotyping, application to larger cohorts, and closing the gap to the rater ceiling through an asymmetric loss that penalizes missed teeth directly rather than correcting the bias after the fact.

The same caveat applies with more force to the derived occlusal measures. They are internally consistent, stable to the loss of a single tooth within the bounds reported in Table S5, and fall in the ranges a clinical examination would give, but not one of them has been compared against an examination of the same mouth, and their agreement with clinical ranges is therefore evidence of plausibility and not of validity. They also inherit the detector’s conservative bias: a measure computed from centroids is only as complete as the detections it is given, and the participants in whom it can be computed at all are, by construction, those with teeth in both jaws. We report them because a segmentation-based pipeline makes them free and because their null association with cognition is itself informative, not because they are ready to be used as dental phenotypes. Anyone wishing to use them that way needs the examination-anchored validation that the tooth count itself still lacks.

## 5. Conclusions

MOLAR turns the dentition captured incidentally in structural brain MRI into a measurable phenotype. Reformulating point-supervised counting as instance segmentation with adjacency-aware capsule labels lets a standard 3D U-Net detect teeth more consistently than raters agree, a learned localizer removes the dependence on registration at a cost of about one tooth, and an image-based usability classifier lets the pipeline decline scans it cannot count. In an independent cohort the automated count reproduced the published association between tooth count and cognition. The measure is calibrated to a rater definition rather than to a dental examination, and anchoring it against examination is the principal next step.

## Supporting information

Supplemental Methods/Results/Discussion

Tables doc

## Code availability

MOLAR will be released open source under an MIT license on acceptance, and is available to reviewers on request, as a pip-installable Python package built on nibabel (Brett et al., 2024), comprising the bite-box pipeline, the automated quality control, the browser annotation tool, and a single command that takes a T1 and returns a bite box, a per-tooth segmentation, tooth counts and QC flags. The ROI backend is swappable: the default localizer has no commercial dependencies, while a registration backend reproduces the reference ROIs exactly for anyone re-deriving the training labels and requires SPM12 Standalone on the free MATLAB Runtime. The two are statistically indistinguishable (paired difference +0.006 teeth, 95% CI −0.264 to +0.276). The backend is recorded in every output record, and the tool refuses by default to write into an output directory produced by a different backend, since mixing backends adds approximately one tooth of noise. Counts use the calibrated threshold by default, arg-max undercounting by roughly 1.6 teeth; the uncalibrated count is also recorded.

## Data availability

The MOLAR pipeline, the annotation tool and the trained weights will be released open source under an MIT license, and the human annotations (per-subject tooth counts, usability ratings and per-tooth marker coordinates) under CC BY 4.0, each archived with a citable DOI. No images are redistributed: dentition is used forensically for identification, so cropped images of a participant’s teeth are not safe to share even though they contain no brain. Anyone holding OASIS-3 access, obtained from its custodians under the OASIS-3 data use agreement, can regenerate every bite box from the released code. The ABC cohort data are clinical and are not released.

## CRediT authorship contribution statement

R.D.N.-N.: Conceptualization, Methodology, Software, Formal analysis, Writing – original draft, Visualization, Writing – review & editing. R.P.: Investigation, Data curation, Writing – original draft, Visualization, Writing – review & editing. S.K.: Data curation, Writing – review & editing. J.S.: Data curation, Writing – review & editing. P.R.K.: Writing – review & editing. S.P.: Writing – review & editing.

## Acknowledgements

Data were provided in part by OASIS-3: Longitudinal Multimodal Neuroimaging: Principal Investigators: T. Benzinger, D. Marcus, J. Morris; NIH P30 AG066444, P50 AG00561, P30 NS09857781, P01 AG026276, P01 AG003991, R01 AG043434, UL1 TR000448, R01 EB009352. The Aging Brain Cohort (ABC) data repository collection was supported by University of South Carolina (USC) Excellence Initiative. RNN is supported by NIH RF1-MH133701 and P50-DC01466.

## Declaration of generative AI and AI-assisted technologies in the writing process

During the preparation of this work the authors used Claude Sonnet 5 to check for grammar issues and revise grammar/sentence structure accordingly. The authors take full responsibility for the content of the published article.

---

**Figure S1:**
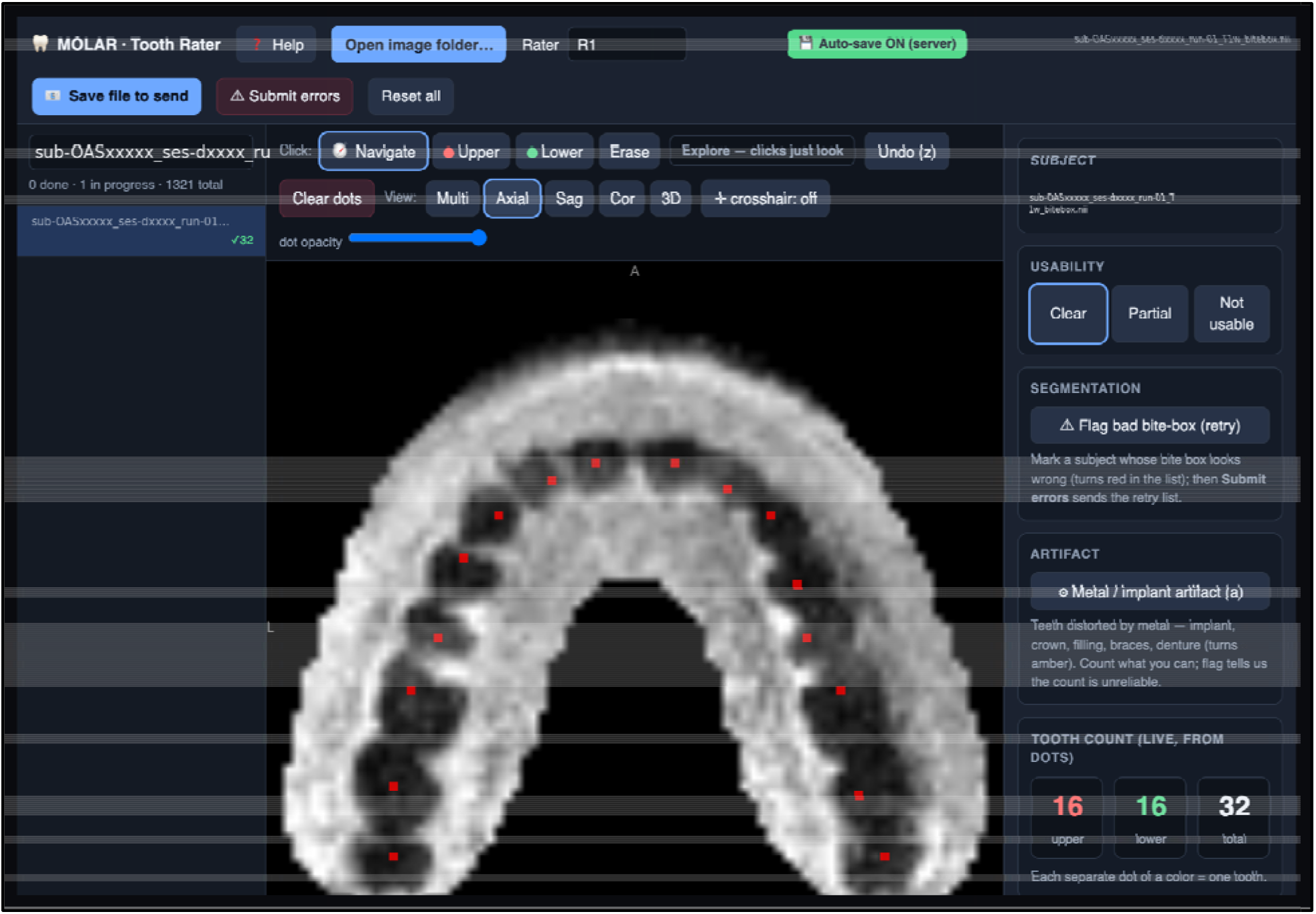
The MOLAR labeling tool. Browser-based annotation interface used to establish ground truth, shown on an upper-arch axial slice with one rater’s markers in place. The rater selects a subject, navigates the bite box, sets a usability rating, flags metal artifact or a bad bite box, and places one marker per tooth for the upper and lower arches; the count updates live as connected components of the markers, and every annotation is saved automatically. Lower-arch markers sit on other slices and are not visible in this view. Participant identifiers have been replaced in the figure, in four places, and no other pixel of the screenshot is altered. The tool is built on the NiiVue WebGL viewer (Hanayik et al. 2026) and runs offline with no installation.

**Figure S2:**
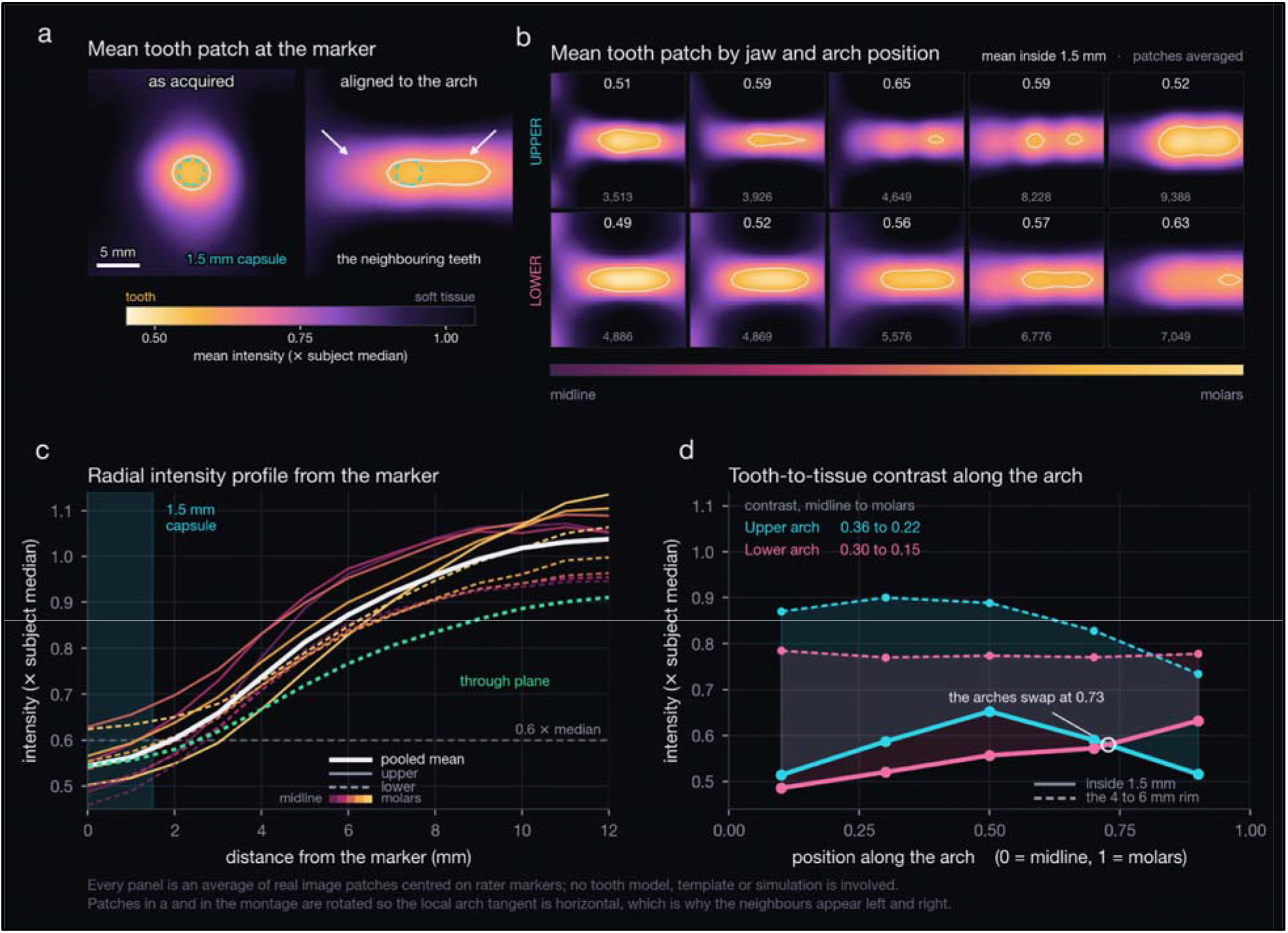
Average tooth appearance at bite-box resolution. Intensity profile around rater markers pooled across the cohort, averaging 6,472 patches centered on markers, showing why teeth are difficult on this modality: the crown is a signal void whose contrast against surrounding structures is low and whose boundary is not sharply defined at 1 mm. The void bottoms out at 0.544 times the subject median at the marker. Beyond about 3.4 mm the average begins to pick up the neighboring tooth’s void, so the outer contour overstates a single tooth, but the 1.5 mm label capsule sits well inside the core.

**Figure S3.**
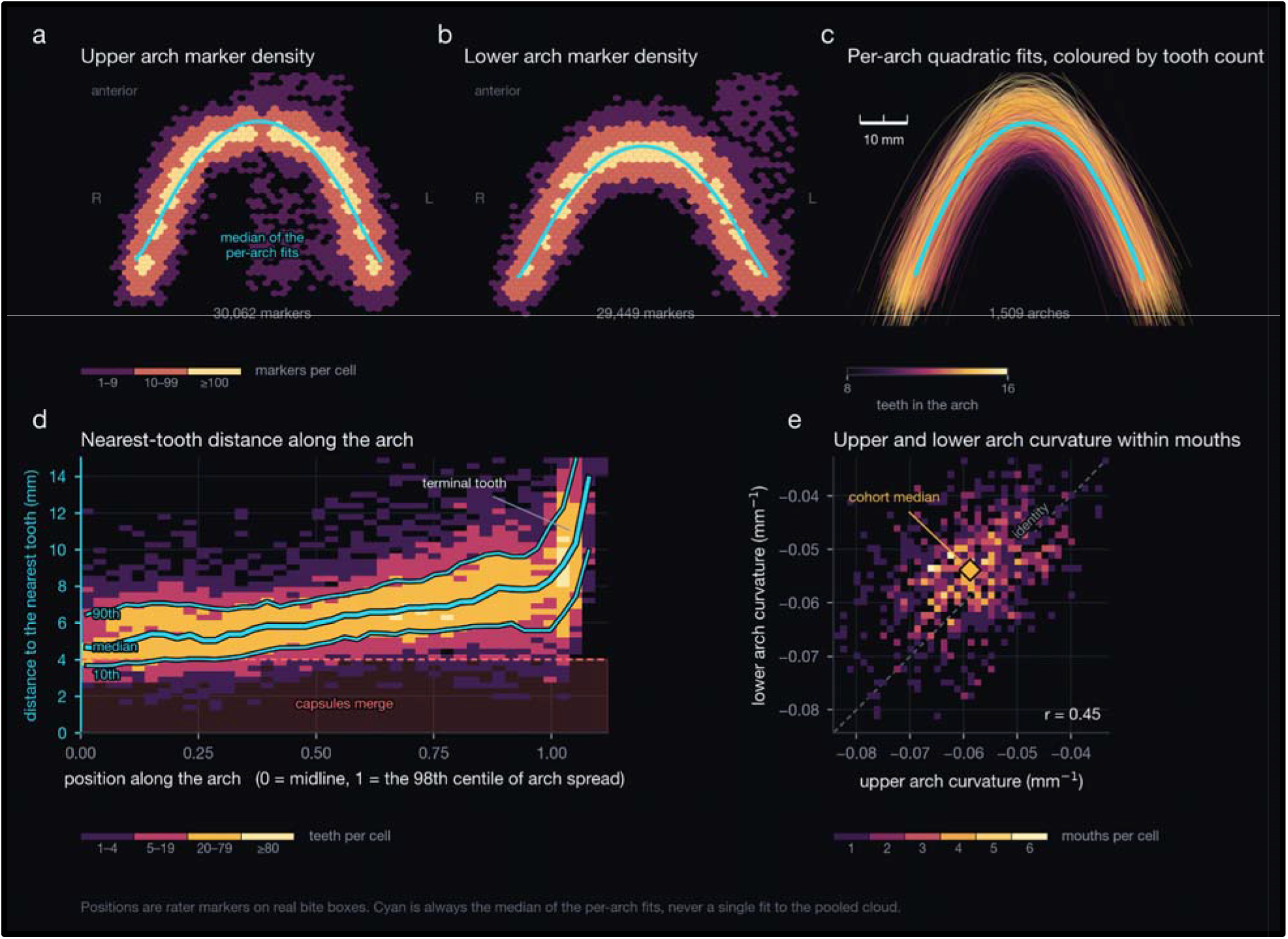
Dental arch geometry across the cohort. Marker positions pooled over all rated subjects, normalized by the bite-box bounding box. Both arches are well described by a downward parabola, fitted curvature −0.052 upper and −0.048 lower, which is what makes tooth identity recoverable from position alone: ordering detections along the left-right axis reproduces the anatomical sequence without any identity labels. Median in-plane nearest-neighbor spacing is 6.57 mm with a 1st percentile of 3.00 mm, the measurement that justifies treating each tooth as a separable connected component.

**Figure S4:**
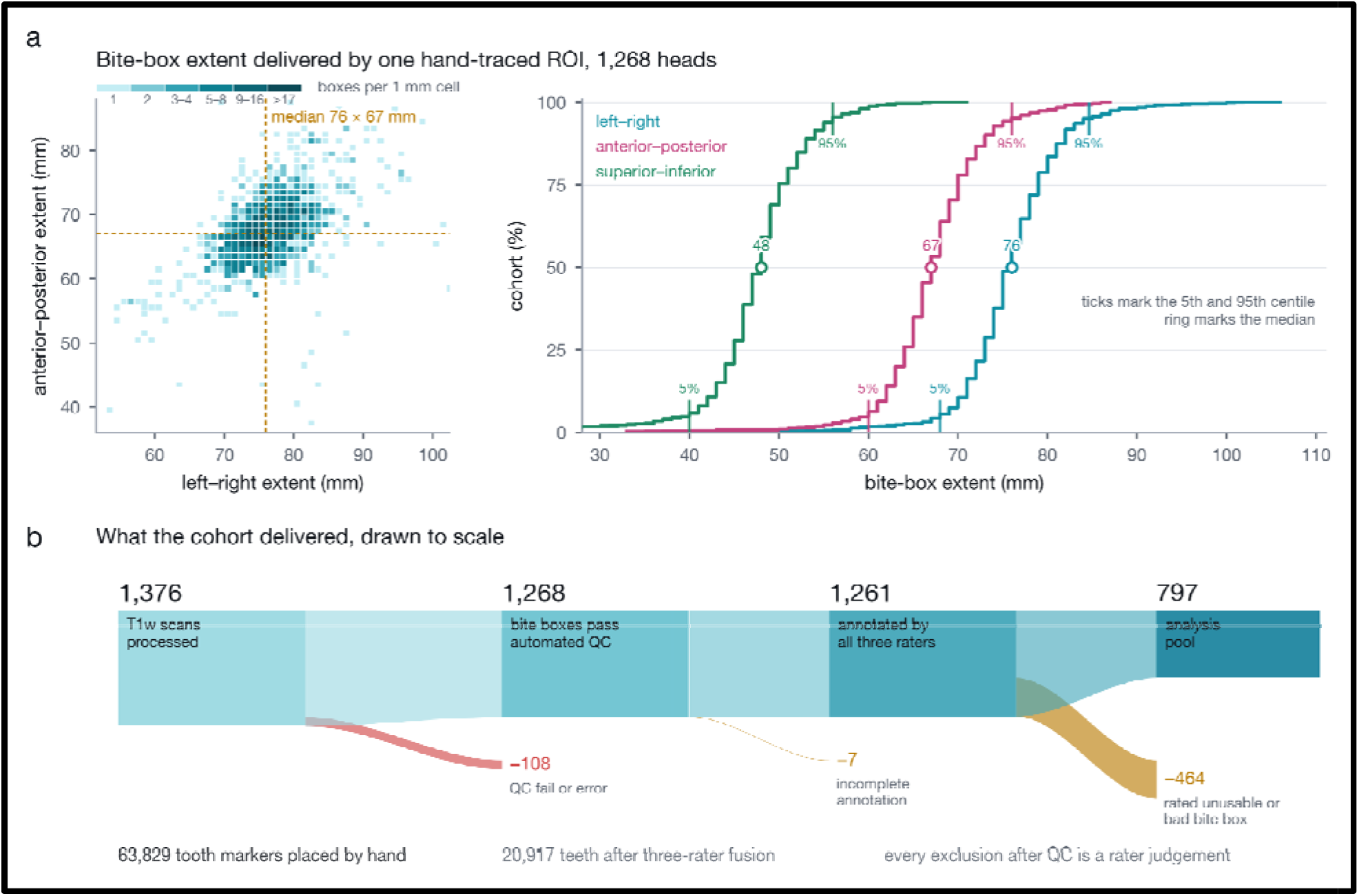
Bite-box geometry and the cohort ledger. a, The geometry the single hand-drawn template tracing delivers, measured on all 1,268 bite boxes the pipeline produced: the joint distribution of left-right against anterior-posterior extent at 1 mm resolution, with the cumulative distribution of each of the three extents beside it. Because the template region is warped to each subject, the extracted volume varies from scan to scan, and this is that variation measured rather than assumed. b, The cohort ledger drawn to one shared scans-per-height scale: 1,376 scans processed, 1,268 passing automated quality control, 1,261 annotated by all three raters and 797 entering the analysis pool. Each loss peels off the underside of the rail at the point it occurs and is labeled with its cause. Every exclusion after quality control is a rater judgment.

**Figure S5:**
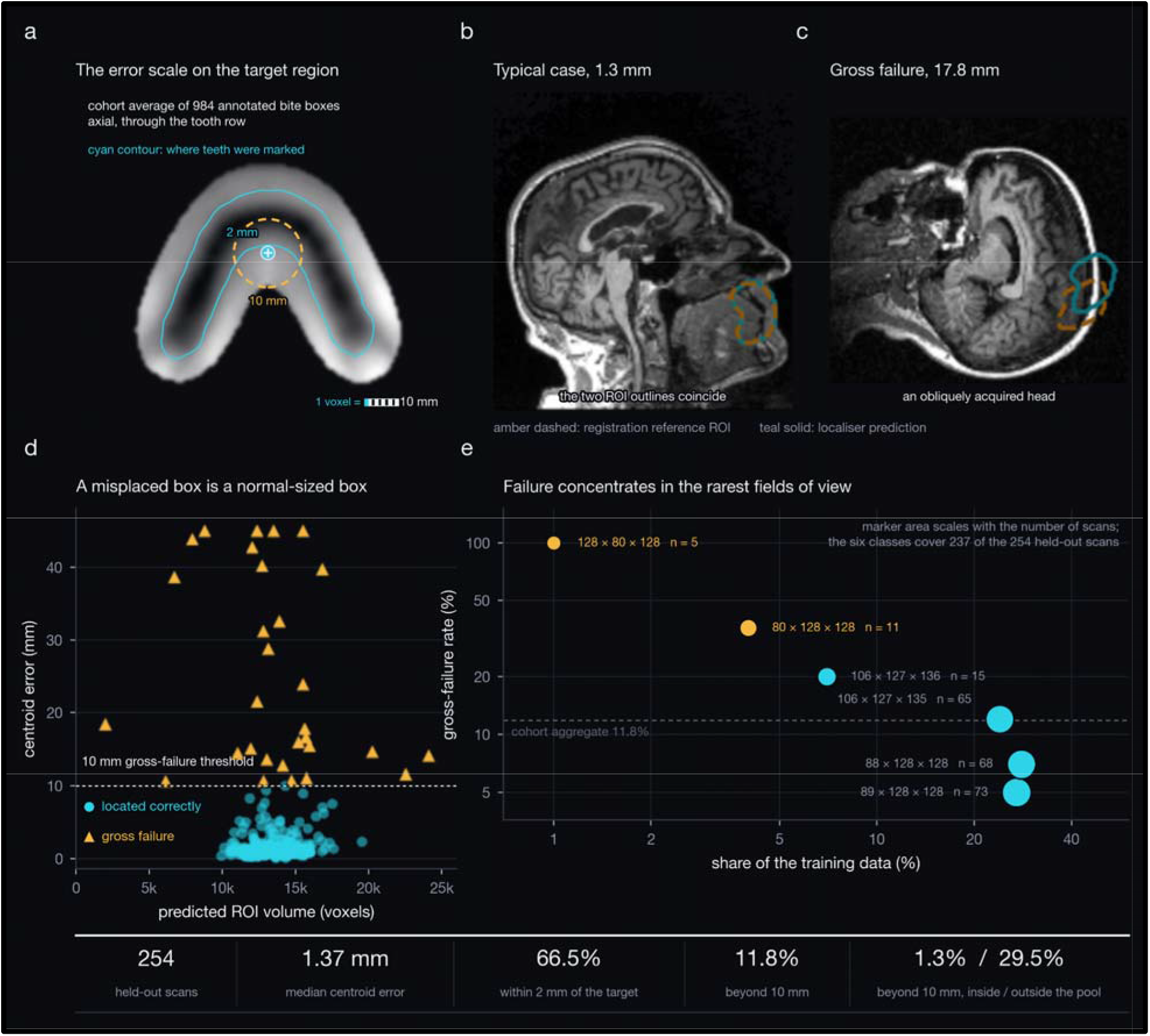
Successes and failures of the localizer. a, The region the localizer is aiming at, with the reported error scale drawn on it. The image is the average of 984 annotated bite boxes resampled onto one 1 mm grid, each box first centered on its own subject’s fused-marker centroid, so the grid center is by construction the point the localizer targets; axial, through the tooth row. The filled disc is the 2 mm radius within which 66.5% of held-out predictions land and the dashed ring is the 10 mm gross-failure threshold, so the median miss is smaller than a tooth while a gross failure is most of a quadrant. The contour is the density of the raters’ own marker coordinates in the same space. b, c, Two held-out examples, a typical case at 1.3 mm and a gross failure at 17.8 mm on an obliquely acquired head; the dashed outline is the registration reference region and the solid outline the localizer prediction. d, Predicted region volume against centroid error: a misplaced box has an entirely normal volume, so a plausibility check on size cannot screen these failures out, which is what motivates flagging by disagreement between two independently trained localizers. e, Gross-failure rate against how rare each input field-of-view shape is in the training data, marker area giving the number of held-out scans carrying that shape; the six shapes shown cover 237 of the 254 held-out scans. Failure concentrates in the rarest geometries, but shape marks truncated acquisitions rather than causing failure, since rebuilding the training set in a fixed 128³ 2 mm cube left the aggregate rate unchanged.

