## Supplemental Tables for "MOLAR: MRI-based Opportunistic Localization and Recognition of teeth"

**Supplementary Tables:**

**Table S1:** Candidate model families and why the three-dimensional U-Net was selected.

| **Family** | **Rationale** | **Decision** |
| --- | --- | --- |
| 3D U-Net, capsules | Points to parts | Selected |
| 3D U-Net, ResEnc-M | Larger encoder | Compared |
| 2D U-Net | Tests 3D context | Compared |
| Count regression | Discards points | Rejected |
| Intensity | No labels needed | Baseline |

3D/2D, three- and two-dimensional. ResEnc-M, residual-encoder medium preset. Points to parts, each tooth represented as a compact capsule on its consensus marker so that the count is the number of 6-connected components. Count regression was rejected rather than tested because it discards the 63,829 point annotations and produces a number that cannot be audited against the image.

**Table S2:** Architecture comparison (majority target, fold 0, identical 160 held-out scans).

| **Config** | **MAE** | **Bias** | **≤ 2** | **F1₃** | **F1₄** |
| --- | --- | --- | --- | --- | --- |
| 3D U-Net | 2.33 | −1.59 | 69.4% | 0.873 | 0.892 |
| ResEnc-M | 2.46 | −1.35 | 61.3% | 0.862 | 0.884 |
| 2D U-Net | 4.41 | +0.01 | 47.5% | 0.786 | 0.811 |
| Human | 1.70 | – | 72-85% | 0.787 | 0.872 |
| Constant | 3.57 | 0.00 | 31.6% | – | – |

3D U-Net, the three-dimensional full-resolution configuration. ResEnc-M, residual-encoder medium preset. MAE, mean absolute error in teeth. Bias, mean signed error. ≤2, proportion of scans within two teeth. F1₃ and F1₄, in-plane detection F1 at 3 mm and 4 mm matching tolerance. Human, the rater-versus-rater ceiling. Constant, always predicting the training-set mean.

**Table S3:** Label-source ablation (fold 0, identical held-out scans, mean absolute error in teeth against each rater's own count).

| **Trained on** | **vs R1** | **vs R2** | **vs R3** | **Mean** | **F1₃** |
| --- | --- | --- | --- | --- | --- |
| 3-rater | 2.93 | 2.19 | 2.71 | 2.61 | 0.880 |
| R1 only | 2.83 | 2.48 | 2.72 | 2.68 | 0.850 |
| R3 only | 3.26 | 2.66 | 2.99 | 2.97 | 0.849 |
| R2 only | 4.39 | 3.31 | 4.15 | 3.95 | 0.843 |
| Rater | 1.64 | 1.87 | 1.60 | 1.70 | 0.787 |

Cells are mean absolute error in teeth against each rater's own count, so no arm is graded by the rater that trained it. 3-rater, trained on the three-rater consensus. R1-R3, the three raters. Rater, the rater-versus-rater ceiling. F1₃, in-plane detection F1 at 3 mm.

**Table S4:** Derived occlusal measures against MoCA (n = 182 ABC participants, partial correlations adjusting for age, sex and race).

| **Measure** | **n** | **Partial r** | **p** | **p, Holm** |
| --- | --- | --- | --- | --- |
| **Tooth count** | 182 | +0.135 | 0.071 | 0.571 |
| **Occluding pairs** | 182 | +0.039 | 0.608 | 1.000 |
| **Overjet** | 171 | +0.046 | 0.553 | 1.000 |
| **Inter-arch distance** | 182 | −0.166 | 0.027 | 0.239 |
| **Arch width** | 182 | +0.080 | 0.291 | 1.000 |
| **Midline deviation absolute** | 182 | −0.066 | 0.384 | 1.000 |
| **Midline offset (signed)** | 182 | −0.087 | 0.249 | 1.000 |
| **Arch curvature, upper** | 182 | +0.025 | 0.743 | 1.000 |
| **Arch curvature, lower** | 182 | −0.036 | 0.629 | 1.000 |

Partial r, correlation with MoCA after removing age, sex and race from both variables. p, Holm, adjusted for the nine measures tested. Sample sizes differ for overjet because it requires anterior teeth in both jaws. The tooth count is shown for reference and is the strongest measure here; no derived measure exceeds it, and none survives correction. The count’s coefficient is lower than the r = +0.177 for the full ABC sample (**Supplementary Results S6**) because the geometric measures require teeth in both jaws, which truncates the tooth-count range in this subset.

**Table S5:** Derived occlusal measures on the ABC cohort (n = 213 with a full measure set).

| **Measure** | **Median** | **IQR** | **Stability** |
| --- | --- | --- | --- |
| **Overjet, mm** | 3.73 | 2.10 to 4.90 | 1.69 |
| **Inter-arch distance, mm** | 14.43 | 13.17 to 16.15 | - |
| **Arch width, upper, mm** | 48.47 | 45.71 to 50.38 | - |
| **Arch width, lower, mm** | 46.52 | 42.60 to 49.82 | - |
| **Midline offset, signed, mm** | 0.11 | −0.71 to 1.33 | 3.09 |
| **Midline offset, absolute, mm** | 1.01 | 0.47 to 1.88 | 3.09 |
| **Occluding pairs, n** | 10 | 8 to 12 | - |
| **Occlusal normal, degrees** | - | - | 0.28 |

IQR, interquartile range. Stability, the worst-case change when any single tooth is dropped, medianed over participants; see Supplementary methods. Occluding pairs, upper teeth with a lower tooth within 5 mm in plane. Midline offset is reported both signed (upper relative to lower) and as its absolute value; **Figure 7b** plots the absolute measure. Overbite is absent from this table because it is not measurable on this modality; inter-arch distance is not a substitute for it.
