## Supplemental Methods/Results/Discussion for "MOLAR: MRI-based Opportunistic Localization and Recognition of teeth"

**Supplementary Methods**

The material below supports design choices described in the Methods but is not required to follow the method or its results.

**Supplementary Methods S1. Bite-box construction and reslicing**

The anchor region of interest is a single expert tracing of all teeth, drawn once on the bone compartment of an extended tissue-probability-map template in MNI space (Fonov et al. 2011). The tracing is never modified; five progressively dilated copies are derived from it, giving six candidate regions of interest of increasing size (TPM_00 through TPM_05; Figure 1d), so that downstream extraction tolerates small registration errors and the appropriate candidate can be selected per participant. Each T1w image is processed in four steps. (1) Tissue segmentation: unified segmentation (SPM12; Ashburner and Friston 2005; Penny et al. 2011) with the extended template priors, retaining the native bone compartment as the registration target. (2) Template-to-native normalization: the template bone compartment is normalized to the participant's own bone compartment with SPM12's normalization routines (Ashburner 2007) and the resulting transform is applied to all six candidate regions, bringing them into native space. (3) Masking: the T1w image is resliced onto each native region grid with trilinear interpolation and intensities are retained only within the region. (4) Cropping: each masked volume is cropped to the tight bounding box of its non-zero voxels, yielding six candidate bite boxes per participant.

The reimplementation was checked against the reference implementation it replaces before any modeling. Every stage downstream of registration reproduced the reference output bit-for-bit on validation cases, and the warped tooth regions and the bone compartment matched at Dice 0.99 and 0.98 to 0.99 respectively. The residual differences were traced to integer rounding at write time rather than to algorithmic divergence and were removed by adopting the reference rounding convention. Every bite box is scored on four measurements: in-region volume, in-region signal fraction, contact with the field-of-view boundary, and the number of large contiguous low-signal clusters. Each scan is assigned ok, review or fail with a categorical reason. Among the excluded OASIS-3 scans the dominant reasons were low in-region signal fraction combined with field-of-view truncation of the dentition (44 scans), an empty or undersized region with low signal (43), and low signal alone (18). Processing took roughly 2.5 min per scan.

**Supplementary Methods S2. Infeasibility of unsupervised dentition localization**

Enamel and dentine are effectively MR-signal-void, so teeth appear as dark structures. Averaging 6,472 image patches centered on rater markers gives a well-defined void reaching approximately 0.6 times the subject-median intensity at the marker and recovering outward (Figure S2). Darkness alone, however, is not discriminative: within a bite box, 98 to 99% of sub-threshold voxels form a single connected component spanning the teeth, the oral cavity and the airway. Intensity thresholding and region growing therefore cannot isolate a tooth. A marker-seeded watershed fails for the same reason. With no background class the seeds must partition the whole connected dark mass, and one basin absorbs it: the median basin size is 1 to 5 voxels per tooth against a single basin exceeding 20,000 voxels. A naive unsupervised dark-blob detector, evaluated as a floor, gave MAE 18.71 teeth with a bias of +18.71, over-detecting for the same reason. Tooth identity must be learned.

**Supplementary Methods S3. Marker geometry and cross-rater marker fusion**

Marker coordinates are reconciled with each image's affine before use, since an annotation tool may report positions in a reoriented frame rather than in on-disk voxel order. Orientation is verified against image content rather than against other labels: teeth are signal voids on T1, so we require that markers fall on darker voxels than their left-right mirror images, which holds in 952 of 975 subjects (paired t = 46.3). Laterality for tooth numbering is derived from the world-x row of the affine, not from its leading diagonal element, because the left-right axis is not the first array axis for 63 of 1,268 bite boxes; the derivation is verified by injecting a marker at a known patient-right coordinate and confirming the assignment across every acquisition orientation present in the cohort. Boxes too oblique for an unambiguous left-right axis (22 of 1,268) are flagged rather than numbered.

Because raters mark the same tooth at slightly different positions and on their own chosen slice, markers are fused before labeling. We anchor on the rater whose count equals the three-rater consensus and refine each position using the nearest marker from each other rater within 3 mm, recording how many raters corroborated each tooth (three raters, 13,673 teeth; two, 5,254; one, 1,990). An initial single-linkage clustering approach was rejected: with a 3 mm link and 6.57 mm median spacing, markers chain across adjacent teeth (rater A's tooth, rater B's inter-tooth marker, rater C's next tooth), silently merging neighbors and losing approximately one tooth per case. Anchoring cannot chain, and it makes the label count equal the consensus count by construction.

**Supplementary Methods S4. Derivation of tooth identity from arch position**

Raters did not label tooth identity, so FDI codes (International Organization for Standardization 2016) are derived rather than supervised. Because each arch is well approximated by a downward parabola (fitted curvature -0.052 upper, -0.048 lower; Figure S3), ordering detections by their left-right coordinate recovers the anatomical sequence directly (Figure 4). Positions are normalized by the bite-box bounding box rather than by the teeth themselves, since centering on the markers would displace the midline whenever posterior teeth are absent, precisely the cases of interest.

Canonical slot positions were estimated from the 257 subjects with a complete 14 + 14 complement, and observed teeth are assigned to slots by a monotone dynamic-programming alignment that permits empty slots, so a partial dentition is numbered with explicit gaps rather than renumbered consecutively. The alignment has no skip-an-observation transition, so arches carrying more observations than there are canonical slots cannot be placed; 105 of 1,594 arches were unplaceable on this criterion and are returned unnumbered. Laterality is taken from the image affine and requires independent confirmation before FDI codes are released for use.

**Supplementary Methods S5. Derived occlusal measures**

The canonical dental frame. Every geometric measure is read in a per-participant frame built from the detections alone, so that it does not depend on how the head was positioned in the scanner. The occlusal plane normal is obtained by fitting a least-squares plane to each jaw’s tooth centroids separately and averaging the two normals after sign-alignment. Fitting both jaws together is the obvious approach and is wrong: the arches sit about 16 mm apart, so a pooled fit describes the slab between them rather than either plane. Measured on this cohort, pooled fits have a residual flatness of 0.424 against 0.007 per jaw, and the pooled normal is a median 36.7 degrees away from the per-jaw one and more than 15 degrees away in 58% of participants. The in-plane axis is set by minimizing the mirror cost of the arch about candidate axes, that is by bilateral symmetry, and the anterior direction is taken as the narrow end of the arch. Volumes are resampled into the frame with trilinear interpolation and labels with nearest-neighbor.

Overjet is a horizontal measure and must be read horizontally. Overjet is computed as the difference between how far forward each arch reaches: for each jaw, the anteroposterior extent of each tooth lying within 10 mm of the front of the arch and 12 mm of the midline is averaged across those teeth, and the lower value is subtracted from the upper. Two details matter. First, an earlier formulation sampled the image intensity along the line between an upper and a lower tooth and took the anteroposterior component of the gap it found; because the jaws are about 15 mm apart vertically that line is nearly vertical, its horizontal component collapses, and the measure returned 0.88 mm at the cohort median and ranked participants incorrectly against visual inspection. A measure of horizontal relationship cannot be derived from a line that spans the jaws. Second, the reach is averaged over teeth rather than read from the single most anterior voxel, which on the 1 mm frame quantizes overjet to whole millimeters, giving 16 distinct values across the cohort against 128 when averaged, and lets one label voxel move the result. The two agree at r = 0.974, so this is a question of precision rather than of construct.

Stability testing. Because these measures are fit to detect centroids rather than to a physical landmark, each was subjected to a leave-one-tooth-out analysis on the 223 participants with at least five teeth in each jaw. Every tooth is dropped in turn, the frame and the measures are recomputed from the remainder, and the change is recorded. Two readings are given: the typical change, the median over drops within a participant, and the worst case, that participant’s largest change; both are then medianed over participants. The typical change is zero for the anterior measures because most drops remove a posterior tooth that does not enter them, so the worst case is the informative figure and is what Table S5 reports.

**Supplementary Results**

**Supplementary Results S1. Assessment of count bias against the training target**

One apparent result is an artifact worth naming, because it is easy to mistake for evidence of robustness. The majority target contains 2.15 fewer teeth per scan than the all-marker target (22.71 against 24.98). A model trained on all markers and scored against the majority target therefore looks unbiased (MAE 1.94, bias -0.09), as though the network had learned to ignore teeth only one rater marked. It has not: trained directly on the majority target, the same undercount reappears (bias -1.59). The apparent agreement is two offsets cancelling, the model's own undercount against the difference between the two label definitions. Bias must be assessed against the target a model was actually trained on.

**Supplementary Results S2. Determinants of localizer failure and detection of gross errors**

Failure rate tracks input field-of-view shape (Figure S5): the three shapes comprising 80% of the training data fail in 5 to 12% of cases, while a shape representing under 1% of the data fails in 5 of 5. Field-of-view shape is a marker of truncated scans rather than the mechanism. Rebuilding the training set in a fixed 128³ 2 mm cube, which removes acquisition geometry as a variable, left the failure rate unchanged (11.8%, with 27 of 30 failures the same scans). Failed predictions land on tissue rather than outside the head (in-region signal 0.998), so the network selects the wrong anatomy rather than diverging, which is consistent with the jaw genuinely being absent from those images.

A second and controllable driver is acquisition obliquity. The pipeline canonicalizes axis order but never removes rotation, so an obliquely acquired or tilted head reaches the network rotated. The scans the localizer mislocates are markedly more oblique than those it gets right (mean 15.2° against 9.7°; 90th percentile 36.2° against 16.6°), and head pitch is twice as variable among them (SD 23.4° against 11.4°). This also explains why fixing the field of view had no effect: a fixed cube containing a rotated head still presents the network with an unseen pose. Rigid alignment to a common frame before localization is the appropriate remedy and is the one outstanding modeling item.

Gross failures can be flagged, though not from a single prediction, since a wrong box has an entirely normal region volume. Two localizers trained independently on different input geometries produce centroids differing by a median of 0.57 mm when both are correct and 2.52 mm when either fails. Combining that disagreement with the image-side quality-control checks catches 73% of gross failures for a 9% false-alarm rate, cutting the residual failure rate among passing scans from 13.0% to 4.3%. The released tool runs both localizers and reports their disagreement per scan rather than silently discarding the scan.

**Supplementary Results S3. Contribution of registration and the sub-voxel phase offset**

With the reference region supplied, the pure-Python implementation of every post-normalization stage reproduced its bite box bit-for-bit (9 of 9 test subjects, maximum voxel difference 0), and the reference arm was byte-identical to the tooth model's training input (20 of 20). Registration's entire contribution to inference is therefore the production of a single binary region mask.

The output grid is further not a template-space reorientation, as the presence of a normalization step might suggest. Across all subjects examined, the write grid is the subject's own voxel lattice, x-flipped and resampled to 1 mm, with a rotation difference of 0.00°. The two grids differ only by a sub-voxel phase offset of 0.02 to 0.46 voxel, varying per subject, inherited from the template bounding box. That offset carries no anatomical information and cannot be reproduced without registration, but a canonical 1 mm grid derived from the T1 alone reproduces the geometry in every other respect, and abandoning the registration grid changes the tooth count by only 0.43 teeth (Table 2, arm A against arm B).

**Supplementary Results S4. Range restriction induced by the usability gate**

Because the pipeline can judge whether a bite box is countable, it is natural to ask whether restricting the analysis to confidently countable scans sharpens the association. It does the opposite. Applying the gate at its cross-validated operating point retains 192 of 233 analyzed scans (82.4%) and removes the association in every arm, the manual counts included: the adjusted partial correlation with MoCA falls to +0.002 for manual (p = 0.98), +0.097 for registration-derived and +0.079 for the fully learned pipeline (n = 159).

The reason is that unusability is not independent of the quantity being measured. Scans the gate declines average 22.3 teeth against 27.0 for those it retains (p = 6 × 10⁻⁵), and the standard deviation of the tooth count falls from 4.36 to 2.97 teeth. Fewer teeth make for a sparser, less tooth-like bite box, so the gate preferentially discards the low end of the exposure and the resulting range restriction attenuates any correlation with it. That the attenuation is just as severe for the manual counts establishes it as a property of the selection rather than of the automated measure. The gate is valuable for what it was built for, telling a user which individual images the tool should not be trusted on, and should not be used to select an analysis sample when tooth count is the exposure of interest. We therefore report the association on the full cohort with localization failures removed, and the gated result as a sensitivity analysis.

**Supplementary Results S5. Duplicated source images in the external cohort**

Three pairs of participant identifiers in the ABC delivery share a byte-identical source image, verified by hash; the duplication is upstream of this work. Within every pair the recorded age differs by between 12 and 41 years and MoCA differs as well, so these are different participants and the image is misfiled for at least one member of each pair. Which member is wrong cannot be recovered from the data, so all six identifiers are excluded rather than one of each pair retained: keeping either would leave a record pairing an image with another participant's age, cognitive score and manual tooth count, which is precisely the pairing every analysis here depends on. The discrepancy has been reported to the cohort custodians. The pairs do provide an unplanned test-retest of the human raters on identical images, on which the manual counts differ by 1, 1 and 4 teeth. The automated count is identical within each pair by construction, the pipeline being deterministic.

**Supplementary Results S6. Beyond the count: derived occlusal geometry**

Beyond the count: geometry the toolbox makes available. Because the model segments teeth individually rather than regressing a number, the arrangement of the dentition is available at no extra cost. We place each participant in a canonical dental frame derived from the detections alone, then read a small set of measures from it (Table S5). The frame is recovered for 227 of the 248 ABC scans; the 21 failures all have fewer than four detected teeth in one jaw, too few to fit a plane, and 213 scans carry the full measure set. Overjet has a median of 3.73 mm with 90% of participants inside the 2 to 8 mm range a clinical examination would call normal to mildly increased, and 7% negative, that is edge-to-edge or reverse. Arch width, inter-arch distance and the count of occluding pairs likewise fall where a dental examination would put them. These are descriptive outputs of the same segmentation, obtained without any additional annotation.

The geometry is stable, but only some of it is. Since every measure is fit to detected centroids, the question is how far one missed or spurious tooth moves it. Dropping each tooth in turn (n = 223 scans with at least five teeth per jaw) leaves the occlusal normal essentially fixed, at 0.03 degrees for a typical drop and 0.28 degrees for the worst drop in a participant. Overjet is unmoved by a typical drop and shifts 1.69 mm in the worst case, roughly half its own median, so it is reportable at the group level but should not be read as a per-participant clinical value. Midline offset was not reportable at all under our first definition, the mean lateral position of the four most anterior teeth, which swings by up to 6.3 mm when one anterior tooth is dropped because a distant tooth is pulled into the average; averaging instead over a fixed 12 mm anterior band, so that the number of contributing teeth is free to vary, halves that to 3.09 mm. We report the band definition and flag the sensitivity, because a measure whose value is smaller than its response to one detection is an artifact of the detector.

One measure is not measurable and should not be attempted. Overbite, the vertical overlap of the incisors, cannot be recovered from a brain-optimized T1. Enamel and the interocclusal space are both signal-void, so there is no edge between the biting surfaces to find: the intensity barrier between the arches reaches only 0.76 times the subject median and clears a 0.8 threshold in just 56% of participants. Three formulations were tried and each returned a quantity that was not overbite, namely the separation of the arches, a value that leaked to zero, and a negative interocclusal gap. We report inter-arch distance, which is well defined, and state plainly that it is not overbite.

None of the derived measures predicts cognition better than the count does. We tested each against MoCA on the ABC participants, with the same covariates and exclusions used above (Table S4). The tooth count remains the strongest single measure. Inter-arch distance is the only measure with an uncorrected p below 0.05, at partial r = −0.166 (p = 0.027), and it does not survive correction for the nine measures tested (Holm p = 0.24). It is also substantially a restatement of tooth loss rather than an independent occlusal finding: it correlates with the count at r = −0.372, and with both in the model neither is significant, the count falling to +0.084 and inter-arch to −0.128, for a gain of 1.2 percentage points of variance over the count alone. The count’s own association is weaker in this subset (r = +0.135, p = 0.071) than in the full sample (r = +0.177, p = 0.015), which is expected rather than anomalous: the geometric measures require teeth in both jaws, so the subset in which they exist excludes the sparse-dentition end of the very range the association depends on, the same restriction documented above for the usability gate. We therefore present these measures as descriptive phenotypes the toolbox makes available, not as validated predictors.
