## Supplementary material for "MOLAR: MRI-based Opportunistic Localization and Recognition of teeth": Tables doc

**Table 1:** The four bite-box arms.

| **Arm** | **ROI source** | **Grid** | **Isolates** |
| --- | --- | --- | --- |
| **A** | Registration | Registration | Reference |
| **B** | Registration | Canonical | Grid and phase |
| **C** | Localizer | Canonical | Fully learned |
| **D** | SimpleITK | Canonical | Alternative |

ROI, region of interest. Canonical, a 1 mm LAS grid derived from the participant's own T1. Reference, the bite box on which the tooth model was trained. Registration, SPM12 unified segmentation with template-to-native normalization. SimpleITK, template-to-subject warping with Mattes mutual information.

**Table 2:** Learned versus registration-based localization (n = 158-159 held-out scans; one tooth model throughout, so every difference is attributable to bite-box construction).

| **Comparison** | **Isolates** | **MAE** | **Bias** | **≤2** |
| --- | --- | --- | --- | --- |
| **A vs B** | Grid and phase | 0.43 | −0.08 | 99.4% |
| **B vs C** | ROI prediction | 0.95 | +0.01 | 91.8% |
| **A vs C** | Fully learned | 1.06 | −0.07 | 89.3% |
| **A vs D** | SimpleITK | 6.37 | +5.39 | 41.1% |
| **Rater vs rater** | Human spread | 1.58 | – | 72-85% |

MAE, mean absolute error in teeth. Bias, mean signed error. ≤ 2, proportion of scans agreeing within two teeth. Arms as defined in **Table 1**. Rater vs rater, the disagreement between two trained human raters on the same scans, given for scale.

**Table 3:** External validation on the ABC cohort (n = 233 analyzed).

| **Measure** | **Manual** | **Registered** | **Learned** |
| --- | --- | --- | --- |
| **MoCA, unadjusted** | +0.414 *** | +0.354 *** | +0.372 *** |
| **MoCA, adjusted** | +0.134 | +0.174 * | +0.170 * |
| **BAG (brainageR), unadjusted** | −0.333 *** | −0.249 *** | −0.240 *** |
| **BAG (brainageR), adjusted** | −0.164 * | −0.188 * | −0.154 * |
| **BAG (DeepBrainNet), unadjusted** | −0.303 *** | −0.195 * | −0.189 * |
| **BAG (DeepBrainNet), adjusted** | −0.062 | −0.088 | −0.080 |
| **MAE vs manual** | 0.72 | 3.56 | 3.16 |
| **r vs manual** | – | 0.654 | 0.707 |
| **Box failures** | – | 1/248 | 0/248 |

Counts are from the orientation-corrected tooth model, five-fold ensembled, applied to bite boxes from each localization route. Of the 247 scans for which a bite box was produced, 8 are excluded because localization failed outright and 6 because they share a duplicated source image with another identifier, leaving 233 analyzed; the covariate set is available for 192 of these. Unadjusted rows are Pearson r; adjusted rows are the partial correlation of tooth count with the outcome from a linear model containing age, sex and race, the covariate set of the previously published manual analysis. Sample sizes: MoCA unadjusted n = 233, adjusted n = 192; BAG (brainageR) adjusted n = 191; BAG (brainageR) unadjusted n = 231; BAG (DeepBrainNet) unadjusted n = 141; BAG (DeepBrainNet) adjusted n = 104. MoCA, Montreal Cognitive Assessment. Abbreviations: BAG, brain-age gap (predicted brain age minus chronological age). MAE, mean absolute error in teeth. Manual, the ABC raters' count; their own rater-versus-rater MAE is 0.72. Box failures, scans for which no bite box was produced, out of the full 248 delivered. Unmarked entries are not significant; * p < 0.05; ** p < 0.01; *** p < 0.001.
